# Infliximab for Treatment of Adults Hospitalized with Moderate or Severe Covid-19

**DOI:** 10.1101/2022.09.22.22280245

**Authors:** Jane A. O’Halloran, Eyal Kedar, Kevin J. Anstrom, Matthew W. McCarthy, Emily R. Ko, Patricia Segura Nunez, Cynthia Boucher, P. Brian Smith, Reynold A. Panettieri, Sabina Mendivil Tuchia de Tai, Martin Maillo, Akram Khan, Alfredo J. Mena Lora, Matthias Salathe, Gerardo Capo, Daniel Rodríguez Gonzalez, Thomas F Patterson, Christopher Palma, Horacio Ariza, Maria Patelli Lima, Anne M. Lachiewicz, John Blamoun, Esteban C. Nannini, Eduardo Sprinz, Analia Mykietiuk, Radica Alicic, Adriana M. Rauseo, Cameron R. Wolfe, Britta Witting, Daniel K. Benjamin, Steven E. McNulty, Pearl Zakroysky, Susan Halabi, Sandra Butler, Jane Atkinson, Stacey J. Adam, Richard Melsheimer, Soju Chang, Lisa LaVange, Michael Proschan, Samuel A. Bozzette, William G. Powderly, ACTIV-1 IM study group members

**Affiliations:** Washington University St. Louis, St Louis, MO; Weill Cornell Medicine in New York; University of North Carolina, Chapel Hill, NC; St. Lawrence Health, Potsdam, NY; Duke University Medical Center, Durham, NC; Hospital Nacional Hipolito Unanue, Lima, Peru; National Center for Advancing Translational Sciences; Duke Clinical Research Institute (DCRI); Robert Wood Johnson Medical School, New Brunswick, NJ; Hospital Central de la Fuerza Aerea del Peru, Lima, Peru; Sanatorio Diagnostico, Santa Fe, Argentina; Oregon Health and Science University, Portland, OR; University of Illinois at Chicago, Chicago, IL; University of Kansas Medical Center, Kansas City, KS; Trinitas Hospital, Elizabeth, NY; Nuevo Hospital Civil de Guadalajara Juan I. Menchaca, Guadalajara, Mexico; University of Texas Health Science Center at San Antonio, San Antonio, TX; University of Rochester School of Medicine and Dentistry, Rochester, NY; Clinica Central S.A., Villa Regina, Argentina; Hospital e Maternidade Celso Pierro - PUC Campinas, Campinas, Brazil; MidMichigan Medical Center – Midland, Midland, MI; Sanatorio Britanico. Santa Fe, Argentina; Hospital de Clinicas de Porto Alegre HCPA, Porto Alegre, Brazil; Instituto Medico Platense, La Plata, Argentina; Providence Medical Research Center, Spokane, WA; Technical Resources International (TRI), Bethesda MD; Foundation for the National Institutes of Health (FNIH); Janssen Pharmaceuticals; National Institute of Allergy and Infectious Diseases (NIAID)

**Keywords:** infliximab, immune modulators, COVID-19, master protocol, shared placebo, TNF alpha inhibitors

## Abstract

**Background:** Immune dysregulation contributes to poorer outcomes in severe Covid-19. Immunomodulators targeting various pathways have improved outcomes. We investigated whether infliximab provides benefit over standard of care.

**Methods:** We conducted a master protocol investigating immunomodulators for potential benefit in treatment of participants hospitalized with Covid-19 pneumonia. We report results for infliximab (single dose infusion) versus shared placebo both with standard of care. Primary outcome was time to recovery by day 29 (28 days after randomization). Key secondary endpoints included 14-day clinical status and 28-day mortality.

**Results:** A total of 1033 participants received study drug (517 infliximab, 516 placebo). Mean age was 54.8 years, 60.3% were male, 48.6% Hispanic or Latino, and 14% Black. No statistically significant difference in the primary endpoint was seen with infliximab compared with placebo (recovery rate ratio 1.13, 95% CI 0.99–1.29; p=0.063). Median (IQR) time to recovery was 8 days (7, 9) for infliximab and 9 days (8, 10) for placebo. Participants assigned to infliximab were more likely to have an improved clinical status at day 14 (OR 1.32, 95% CI 1.05–1.66). Twenty-eight-day mortality was 10.1% with infliximab versus 14.5% with placebo, with 41% lower odds of dying in those receiving infliximab (OR 0.59, 95% CI 0.39–0.90). No differences in risk of serious adverse events including secondary infections.

**Conclusions:** Infliximab did not demonstrate statistically significant improvement in time to recovery. It was associated with improved 14-day clinical status and substantial reduction in 28- day mortality compared with standard of care.

**Trial registration:** ClinicalTrials.gov (NCT04593940).

## INTRODUCTION

Immune dysregulation induced by severe acute respiratory syndrome coronavirus-2 (SARS-CoV-2) is a major cause of morbidity and mortality.^1^ While treatments directly targeting SARS-CoV-2 have shown significant impact in earlier stages of Covid-19,^2^ immunoodulating agents provide benefit in hypoxia observed in later disease stages.^3-6^

Tumor necrosis factor alpha (TNF) is a pro-inflammatory cytokine that plays a role in nearly all acute inflammatory reactions.^7^ In patients hospitalized for Covid-19, increased TNF levels are associated with more severe disease and death.^8^ Inhibition of TNF reduces disease severity in mouse models of other respiratory viruses.^9^ Infliximab, a TNF inhibitor that binds both soluble and transmembrane forms of TNF, is approved and commonly used to treat autoimmune diseases, including inflammatory bowel disease and rheumatoid arthritis.

Data on infliximab use in Covid-19 treatment is limited.^10-12^ However, patients who developed Covid-19 while on TNF inhibitors for other indications did not experience adverse outcomes.^13,14^

In April 2020, the National Institutes of Health (NIH) launched a public-private partnership, Accelerating Covid-19 Therapeutic Interventions and Vaccines (ACTIV), to develop a coordinated research response to Covid-19. ACTIV-1 IM was a master protocol designed to evaluate immunomodulatory agents in hospitalized patients with moderate/severe Covid-19.

Infliximab was one of the agents included in ACTIV-1 IM based on its mechanism of action and efficacy and safety profile in inflammatory disorders. We report the results of a randomized, double-blind, placebo-controlled evaluation of infliximab compared with placebo in addition to standard of care.

## METHODS

### Study design

The ACTIV-1 IM master protocol was developed to allow for parallel investigation of efficacy and safety of multiple immunomodulators compared with a shared placebo, with standard of care given as background therapy in both arms. Remdesivir (Gilead Sciences, Foster City, CA) was provided as standard of care and given to eligible participants.

### Eligibility

Eligibility criteria are outlined in the **Supplementary Appendix**. Briefly, adults ≥18 years with confirmed moderate/severe SARS-CoV-2 infection admitted to hospital were eligible.

Radiological evidence of pulmonary involvement or oxygen saturation ≤94% on room air or supplemental oxygen, mechanical ventilation, or extracorporeal membrane oxygenation (ECMO) was required. Exclusion criteria included aspartate aminotransferase or alanine aminotransferase >10 times the upper limit of normal, estimated glomerular filtration rate <30 mL/min, history of New York Heart Association class III/IV congestive heart failure, neutropenia, lymphopenia, targeted immune therapies for any indication in the last four weeks or five drug half-lives, or evidence of untreated tuberculosis or other untreated infections. Participants were excluded from the infliximab substudy if they had a history of hepatosplenic T-cell lymphoma or other lymphoma within five years before screening, history of or current diagnosis of multiple sclerosis or other significant demyelinating condition.

Eligible participants were randomized in a two-stage process (**Supplementary Appendix**). Between October 16, 2020, and December 31, 2021, 1061 participants were randomized at 69 sites across five countries. Infliximab was administered on day 1 as a single-dose intravenous infusion of 5 mg/kg over at least 2 hours. All participants received local standard of care.

### Procedures

Participants’ clinical status was captured daily through day 29 if hospitalized. For discharged participants, clinical status was assessed on days 8, 11, 15, and 29 in-person or by telephone if in-person assessment was not possible. Day 60 follow-up was conducted by telephone (**Supplementary Appendix**).

### Outcomes and statistical analysis

The primary outcome was time to recovery evaluated up to day 29 (for clinical status on day 28). Recovery was defined as the first day on which participants attained category 6, 7, or 8 on the 8-point ordinal scale (OS) defined in **Supplementary Appendix**. The primary efficacy analysis was based on the Fine-Gray model with stratification by region and baseline disease severity.^15^

Key secondary outcomes were mortality and clinical status assessed by 8-point OS at days 14 and 28. Logistic and ordinal logistic regression models were used to estimate treatment effects for mortality and 8-point OS endpoints. A multiple imputation approach was used to account for the small amount of missing data for key secondary endpoints (**Statistical Analysis Plan [SAP]**). The gatekeeping approach for controlling Type I error for the primary endpoint, day 14 clinical status, and day 28 mortality is described in the SAP, including relevant p-value cutoffs (**Table S1**).

Safety assessments included a composite endpoint of death, serious adverse events (SAEs), or grade 3 (severe) and 4 (potentially life-threatening) AEs occurring through day 60. Secondary infections as AEs of special interest through day 60 and discontinuation or temporary suspension of trial-product administration for any reason were also assessed.

All efficacy and safety analyses reported are based on the modified intention-to-treat (mITT) population consisting of all randomized participants who received at least one dose of assigned study drug (infliximab or shared placebo), limiting shared placebo participants to those eligible for infliximab.

## RESULTS

### Participants

Of 1061 participants who underwent randomization in this substudy, 531 were assigned to the infliximab group and 530 to the shared placebo group (**Figure S1**). Ultimately, 517 participants in the infliximab group and 516 in the shared placebo group received at least one dose of assigned treatment and constitute the mITT population. Consistent with the protocol-specified mITT definition, four participants were excluded post-randomization because they received incorrect study drug. At baseline, 578 (56.0%) participants had moderate disease (52.1% and 3.9% OS 4 and 5) and 455 (44.0%) had severe disease (10.7% and 33.3% OS 2 and 3). Study discontinuation by day 29 occurred in 4.8% in the infliximab group and 4.5% in the shared placebo group. Mean age was 54.8 years and 60% were male. Overall, 652 (63.1%) were White, 145 (14.0%) Black, 27 (2.6%) Asian, and 9 (0.9%) American Indian or Alaska Native; 502 (48.6%) were Hispanic or Latino. Overall, 975 (94.4%) participants received remdesivir, while 950 (92.0%) received corticosteroids (**Table 1**). Characteristics by region are shown in **Table S2, S3**.

**Table 1.**
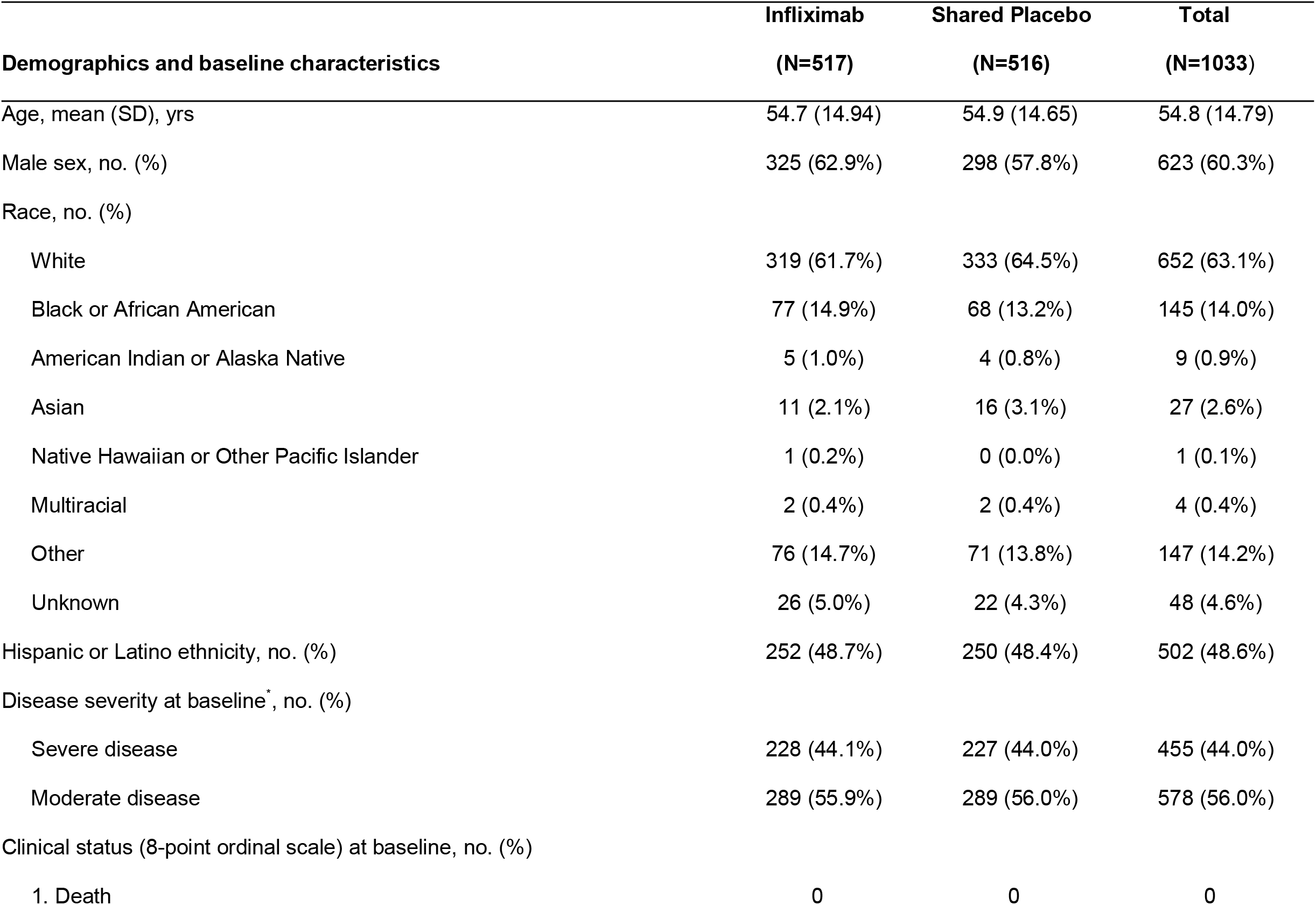
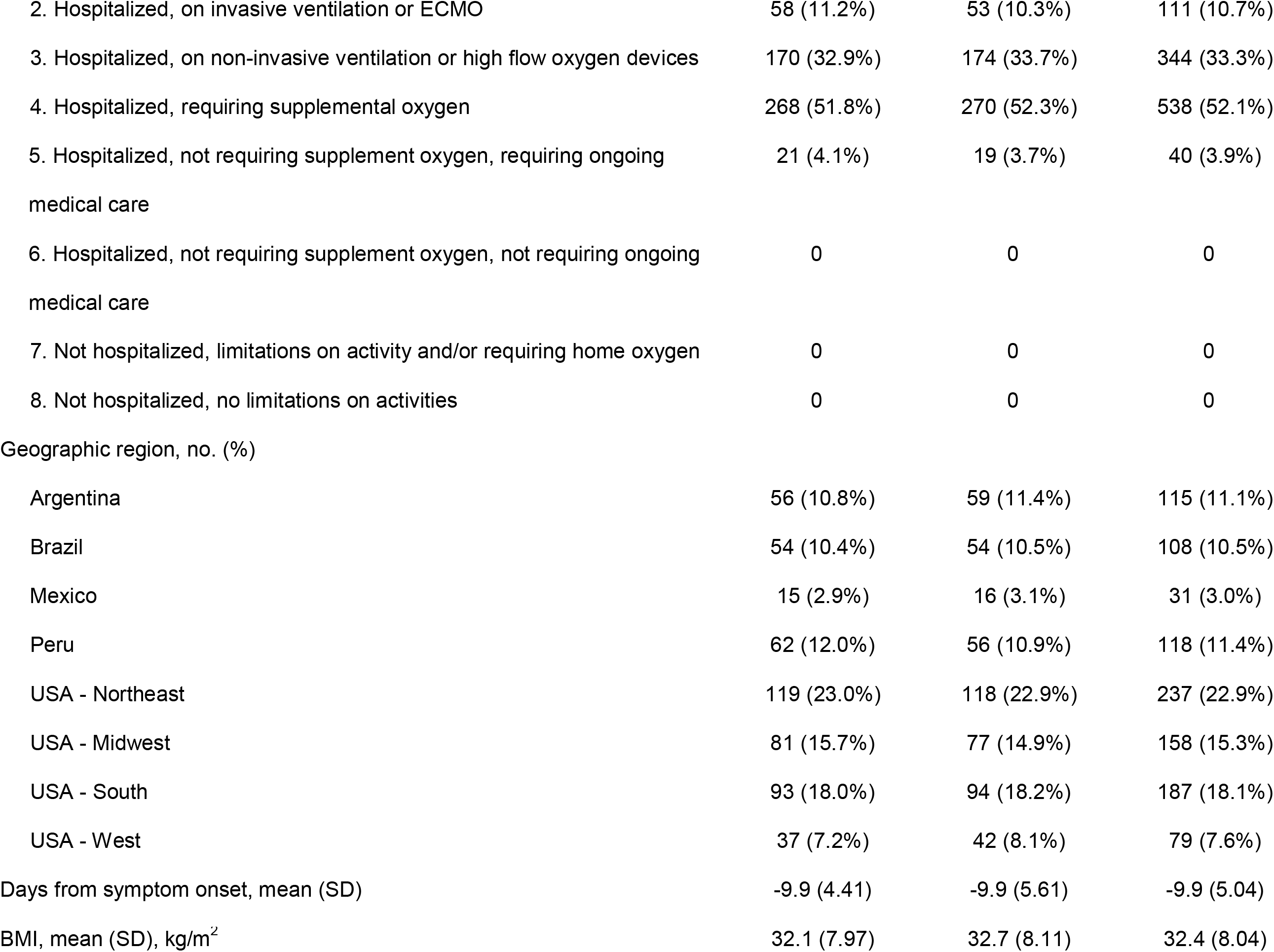
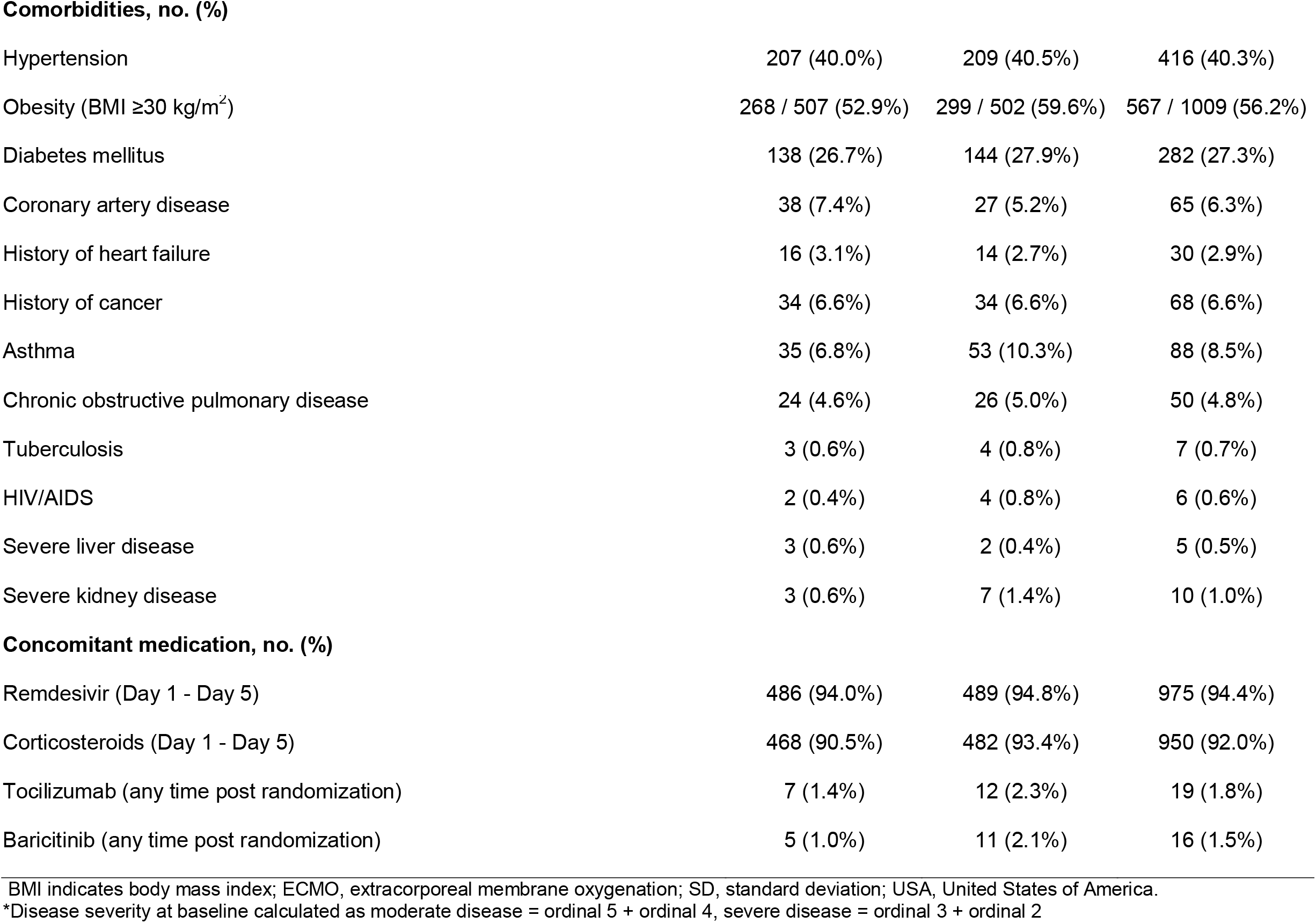
Demographics and baseline characteristics (modified intent-to-treat)

|  | **Infliximab** | **Shared Placebo** | **Total** |
| --- | --- | --- | --- |
| **Demographics and baseline characteristics** | **(N=517)** | **(N=516)** | **(N=1033)** |
| Age, mean (SD), yrs | 54.7 (14.94) | 54.9 (14.65) | 54.8 (14.79) |
| Male sex, no. (%) | 325 (62.9%) | 298 (57.8%) | 623 (60.3%) |
| Race, no. (%) |  |  |  |
| White | 319 (61.7%) | 333 (64.5%) | 652 (63.1%) |
| Black or African American | 77 (14.9%) | 68 (13.2%) | 145 (14.0%) |
| American Indian or Alaska Native | 5 (1.0%) | 4 (0.8%) | 9 (0.9%) |
| Asian | 11 (2.1%) | 16 (3.1%) | 27 (2.6%) |
| Native Hawaiian or Other Pacific Islander | 1 (0.2%) | 0 (0.0%) | 1 (0.1%) |
| Multiracial | 2 (0.4%) | 2 (0.4%) | 4 (0.4%) |
| Other | 76 (14.7%) | 71 (13.8%) | 147 (14.2%) |
| Unknown | 26 (5.0%) | 22 (4.3%) | 48 (4.6%) |
| Hispanic or Latino ethnicity, no. (%) | 252 (48.7%) | 250 (48.4%) | 502 (48.6%) |
| Disease severity at baseline*, no. (%) |  |  |  |
| Severe disease | 228 (44.1%) | 227 (44.0%) | 455 (44.0%) |
| Moderate disease | 289 (55.9%) | 289 (56.0%) | 578 (56.0%) |
| Clinical status (8-point ordinal scale) at baseline, no. (%) |  |  |  |
| 1. Death | 0 | 0 | 0 |
| 2. Hospitalized, on invasive ventilation or ECMO | 58 (11.2%) | 53 (10.3%) | 111 (10.7%) |
| 3. Hospitalized, on non-invasive ventilation or high flow oxygen devices | 170 (32.9%) | 174 (33.7%) | 344 (33.3%) |
| 4. Hospitalized, requiring supplemental oxygen | 268 (51.8%) | 270 (52.3%) | 538 (52.1%) |
| 5. Hospitalized, not requiring supplement oxygen, requiring ongoing medical care | 21 (4.1%) | 19 (3.7%) | 40 (3.9%) |
| 6. Hospitalized, not requiring supplement oxygen, not requiring ongoing medical care | 0 | 0 | 0 |
| 7. Not hospitalized, limitations on activity and/or requiring home oxygen | 0 | 0 | 0 |
| 8. Not hospitalized, no limitations on activities | 0 | 0 | 0 |
| Geographic region, no. (%) |  |  |  |
| Argentina | 56 (10.8%) | 59 (11.4%) | 115 (11.1%) |
| Brazil | 54 (10.4%) | 54 (10.5%) | 108 (10.5%) |
| Mexico | 15 (2.9%) | 16 (3.1%) | 31 (3.0%) |
| Peru | 62 (12.0%) | 56 (10.9%) | 118 (11.4%) |
| USA - Northeast | 119 (23.0%) | 118 (22.9%) | 237 (22.9%) |
| USA - Midwest | 81 (15.7%) | 77 (14.9%) | 158 (15.3%) |
| USA - South | 93 (18.0%) | 94 (18.2%) | 187 (18.1%) |
| USA - West | 37 (7.2%) | 42 (8.1%) | 79 (7.6%) |
| Days from symptom onset, mean (SD) | -9.9 (4.41) | -9.9 (5.61) | -9.9 (5.04) |
| BMI, mean (SD), kg/m<sup>2</sup> | 32.1 (7.97) | 32.7 (8.11) | 32.4 (8.04) |
| **Comorbidities, no. (%)** |  |  |  |
| Hypertension | 207 (40.0%) | 209 (40.5%) | 416 (40.3%) |
| Obesity (BMI $\geq 30$ kg/m<sup>2</sup>) | 268 / 507 (52.9%) | 299 / 502 (59.6%) | 567 / 1009 (56.2%) |
| Diabetes mellitus | 138 (26.7%) | 144 (27.9%) | 282 (27.3%) |
| Coronary artery disease | 38 (7.4%) | 27 (5.2%) | 65 (6.3%) |
| History of heart failure | 16 (3.1%) | 14 (2.7%) | 30 (2.9%) |
| History of cancer | 34 (6.6%) | 34 (6.6%) | 68 (6.6%) |
| Asthma | 35 (6.8%) | 53 (10.3%) | 88 (8.5%) |
| Chronic obstructive pulmonary disease | 24 (4.6%) | 26 (5.0%) | 50 (4.8%) |
| Tuberculosis | 3 (0.6%) | 4 (0.8%) | 7 (0.7%) |
| HIV/AIDS | 2 (0.4%) | 4 (0.8%) | 6 (0.6%) |
| Severe liver disease | 3 (0.6%) | 2 (0.4%) | 5 (0.5%) |
| Severe kidney disease | 3 (0.6%) | 7 (1.4%) | 10 (1.0%) |
| **Concomitant medication, no. (%)** |  |  |  |
| Remdesivir (Day 1 - Day 5) | 486 (94.0%) | 489 (94.8%) | 975 (94.4%) |
| Corticosteroids (Day 1 - Day 5) | 468 (90.5%) | 482 (93.4%) | 950 (92.0%) |
| Tocilizumab (any time post randomization) | 7 (1.4%) | 12 (2.3%) | 19 (1.8%) |
| Baricitinib (any time post randomization) | 5 (1.0%) | 11 (2.1%) | 16 (1.5%) |

BMI indicates body mass index; ECMO, extracorporeal membrane oxygenation; SD, standard deviation; USA, United States of America.
\*Disease severity at baseline calculated as moderate disease = ordinal 5 + ordinal 4, severe disease = ordinal 3 + ordinal 2

### Primary outcome

Median time to recovery for infliximab was 1 day shorter than with shared placebo, but the difference did not reach statistical significance (median 8 vs. 9 days; recovery rate ratio [RRR] 1.13, 95% confidence interval [CI] 0.99–1.29; p=0.063) (**Table 2, Figure S2**). Across OS subgroups, the interaction p-value was 0.36 indicating no difference. Median time to recovery among participants receiving mechanical ventilation/ECMO at enrollment (OS 2) was 23 days for infliximab and >28 for shared placebo (RRR 1.117, 95% CI 0.612–2.039). For those on noninvasive ventilation or high-flow oxygen (OS 3), median time to recovery was 11 days for infliximab and 13 for shared placebo (RRR 1.326, 95% CI 1.039–1.693). Among those hospitalized requiring supplemental oxygen (OS 4) and those hospitalized not requiring oxygen (OS 5), median time to recovery was 6 versus 7 days (RRR 1.090, 95% CI 0.923–1.286) and 5 versus 4 days (RRR 0.767, 95% CI 0.404–1.455) (**Table 2**).

**Table 2.**
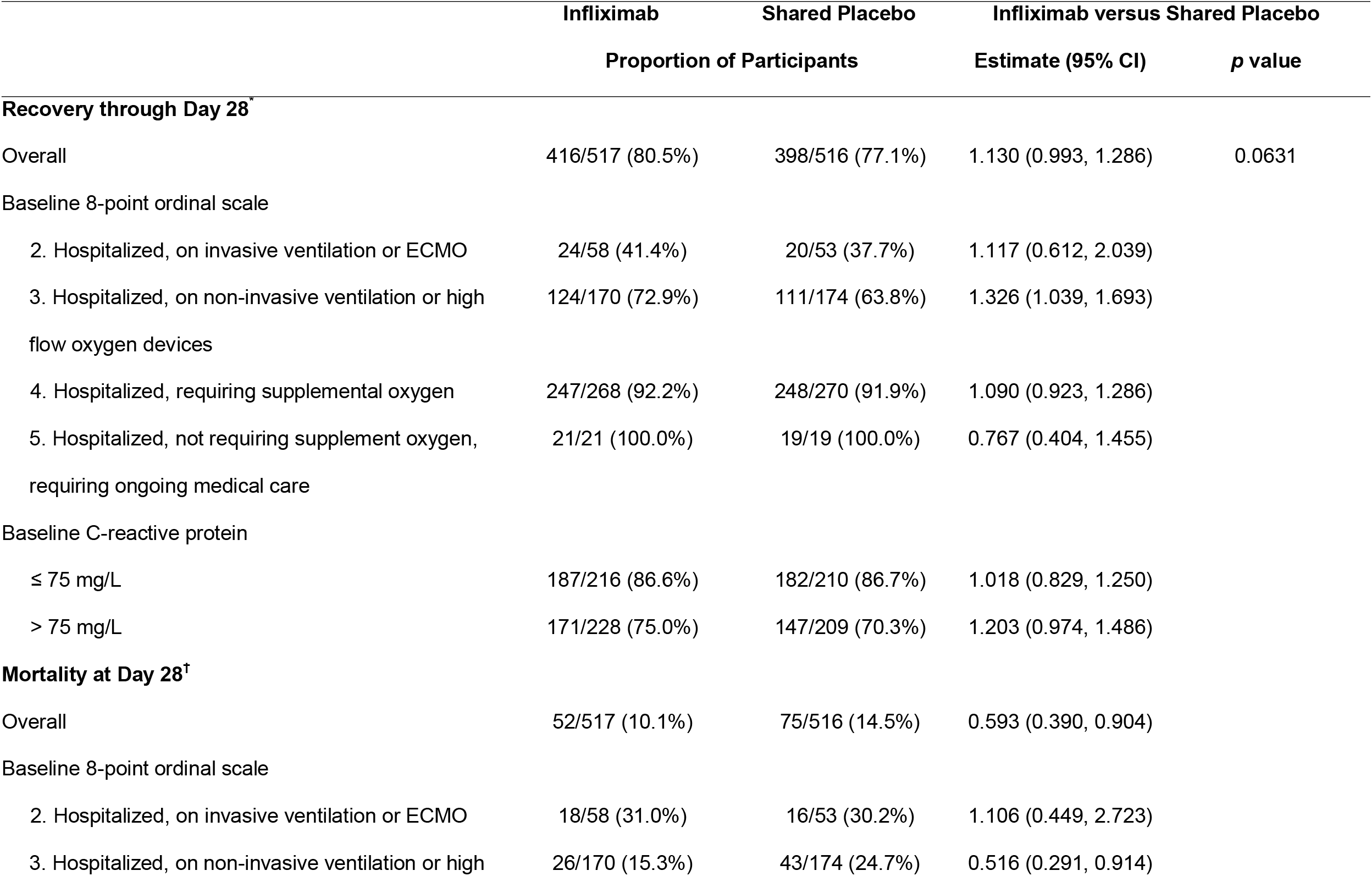
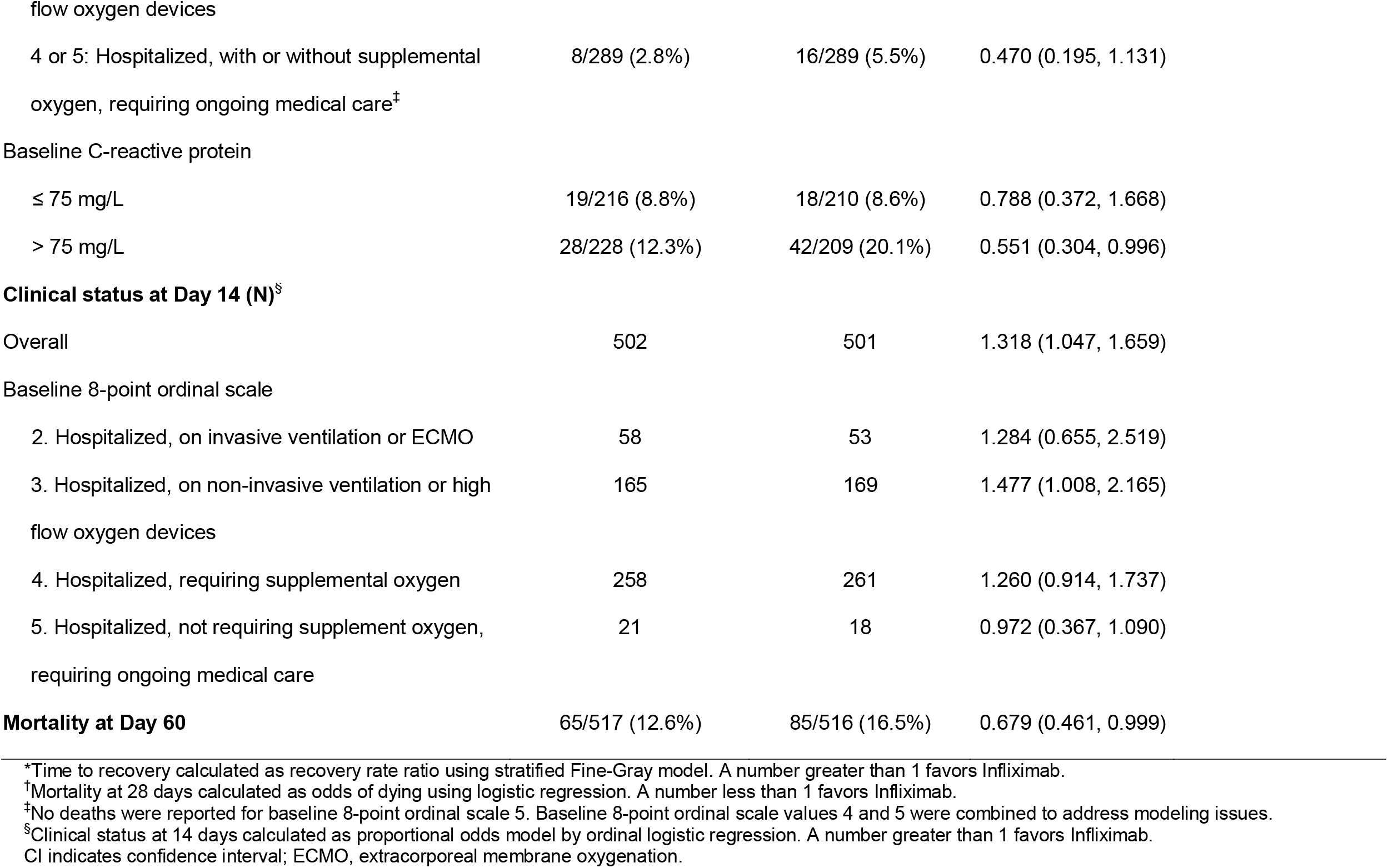
Primary and Key Secondary Endpoints (Modified Intent-to-Treat Population)

|  | Infliximab | Shared Placebo | Infliximab versus Shared Placebo |  |
| --- | --- | --- | --- | --- |
|  | Proportion of Participants |  | Estimate (95% CI) | *p* value |
| **Recovery through Day 28<sup>†</sup>** |  |  |  |  |
| Overall | 416/517 (80.5%) | 398/516 (77.1%) | 1.130 (0.993, 1.286) | 0.0631 |
| Baseline 8-point ordinal scale |  |  |  |  |
| 2. Hospitalized, on invasive ventilation or ECMO | 24/58 (41.4%) | 20/53 (37.7%) | 1.117 (0.612, 2.039) |  |
| 3. Hospitalized, on non-invasive ventilation or high flow oxygen devices | 124/170 (72.9%) | 111/174 (63.8%) | 1.326 (1.039, 1.693) |  |
| 4. Hospitalized, requiring supplemental oxygen | 247/268 (92.2%) | 248/270 (91.9%) | 1.090 (0.923, 1.286) |  |
| 5. Hospitalized, not requiring supplement oxygen, requiring ongoing medical care | 21/21 (100.0%) | 19/19 (100.0%) | 0.767 (0.404, 1.455) |  |
| Baseline C-reactive protein |  |  |  |  |
| ≤ 75 mg/L | 187/216 (86.6%) | 182/210 (86.7%) | 1.018 (0.829, 1.250) |  |
| > 75 mg/L | 171/228 (75.0%) | 147/209 (70.3%) | 1.203 (0.974, 1.486) |  |
| **Mortality at Day 28<sup>†</sup>** |  |  |  |  |
| Overall | 52/517 (10.1%) | 75/516 (14.5%) | 0.593 (0.390, 0.904) |  |
| Baseline 8-point ordinal scale |  |  |  |  |
| 2. Hospitalized, on invasive ventilation or ECMO | 18/58 (31.0%) | 16/53 (30.2%) | 1.106 (0.449, 2.723) |  |
| 3. Hospitalized, on non-invasive ventilation or high | 26/170 (15.3%) | 43/174 (24.7%) | 0.516 (0.291, 0.914) |  |

|  |  |  |  |
| --- | --- | --- | --- |
| flow oxygen devices |  |  |  |
| 4 or 5: Hospitalized, with or without supplemental oxygen, requiring ongoing medical care<sup>†</sup> | 8/289 (2.8%) | 16/289 (5.5%) | 0.470 (0.195, 1.131) |
| Baseline C-reactive protein |  |  |  |
| ≤ 75 mg/L | 19/216 (8.8%) | 18/210 (8.6%) | 0.788 (0.372, 1.668) |
| > 75 mg/L | 28/228 (12.3%) | 42/209 (20.1%) | 0.551 (0.304, 0.996) |
| **Clinical status at Day 14 (N)<sup>§</sup>** |  |  |  |
| Overall | 502 | 501 | 1.318 (1.047, 1.659) |
| Baseline 8-point ordinal scale |  |  |  |
| 2. Hospitalized, on invasive ventilation or ECMO | 58 | 53 | 1.284 (0.655, 2.519) |
| 3. Hospitalized, on non-invasive ventilation or high flow oxygen devices | 165 | 169 | 1.477 (1.008, 2.165) |
| 4. Hospitalized, requiring supplemental oxygen | 258 | 261 | 1.260 (0.914, 1.737) |
| 5. Hospitalized, not requiring supplement oxygen, requiring ongoing medical care | 21 | 18 | 0.972 (0.367, 1.090) |
| **Mortality at Day 60** | 65/517 (12.6%) | 85/516 (16.5%) | 0.679 (0.461, 0.999) |

\*Time to recovery calculated as recovery rate ratio using stratified Fine-Gray model. A number greater than 1 favors Infliximab.
<sup>†</sup>Mortality at 28 days calculated as odds of dying using logistic regression. A number less than 1 favors Infliximab.
<sup>‡</sup>No deaths were reported for baseline 8-point ordinal scale 5. Baseline 8-point ordinal scale values 4 and 5 were combined to address modeling issues.
<sup>§</sup>Clinical status at 14 days calculated as proportional odds model by ordinal logistic regression. A number greater than 1 favors Infliximab.
CI indicates confidence interval; ECMO, extracorporeal membrane oxygenation.

### Key secondary outcomes

#### Mortality

Mortality at day 28 was 10.1% for infliximab and 14.5% for shared placebo (odds ratio [OR] for death 0.59, 95% CI 0.39–0.90), resulting in 41% lower adjusted odds of dying (**Table 2, Figures 1, 2A**). When 28-day mortality was examined by OS, the interaction p-value was 0.31 indicating no difference across subgroups. No difference in mortality was observed in the most severe disease (OS 2) (OR 1.11, 95% CI 0.45–2.72) (**Figure 2C**). However, mortality decreased in those who received infliximab compared with placebo for OS 3 (OR 0.52, 95% CI 0.29–0.91) and OS 4/5 (OR 0.47, 95% CI 0.20–1.13). (**Figure 2D, E**) No deaths occurred in the OS 5 group. Day 14 mortality was 5.6% for infliximab and 8.1% for shared placebo (OR 0.63, 95% CI 0.36–1.08). Additional analysis revealed 60-day mortality rates of 12.6% in the infliximab group and 16.5% in the shared placebo group, with a 32% reduction of odds of mortality observed (OR 0.68, 95% CI 0.46–0.999). The vast majority of those who died between day 29 and 60 were intubated at day 28. In subgroup analyses, although point estimates indicated benefit for infliximab in both c-reactive protein (CRP) subgroups, there was a trend for a stronger mortality reduction at 28 days in participants with CRP >75 mg/L at baseline compared with CRP ≤75 mg/L.

**Figure 1.**
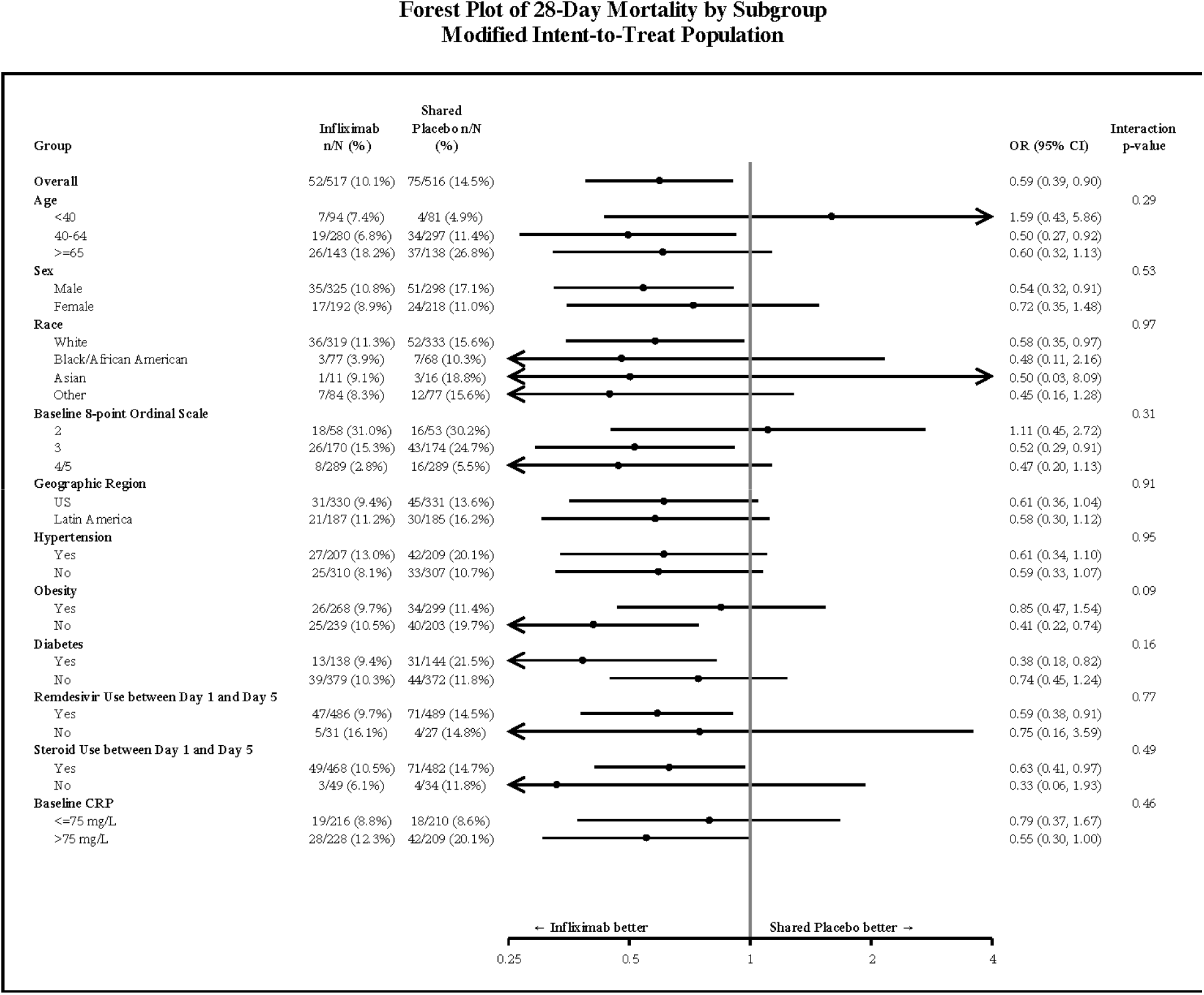
Forest plot of 28-day mortality by subgroup (modified intent-to-treat population) **Figure 2**.

**Figure 2.**
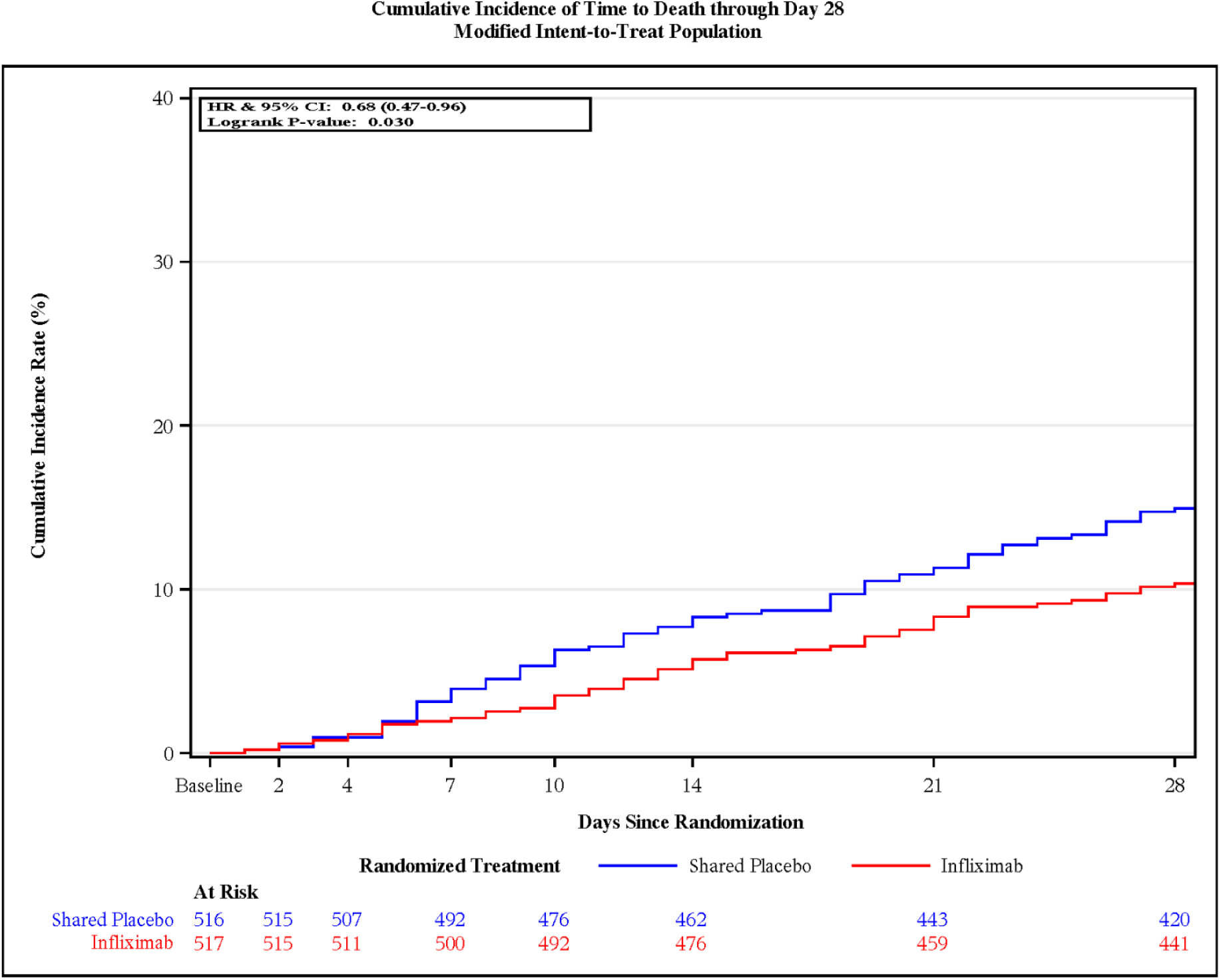
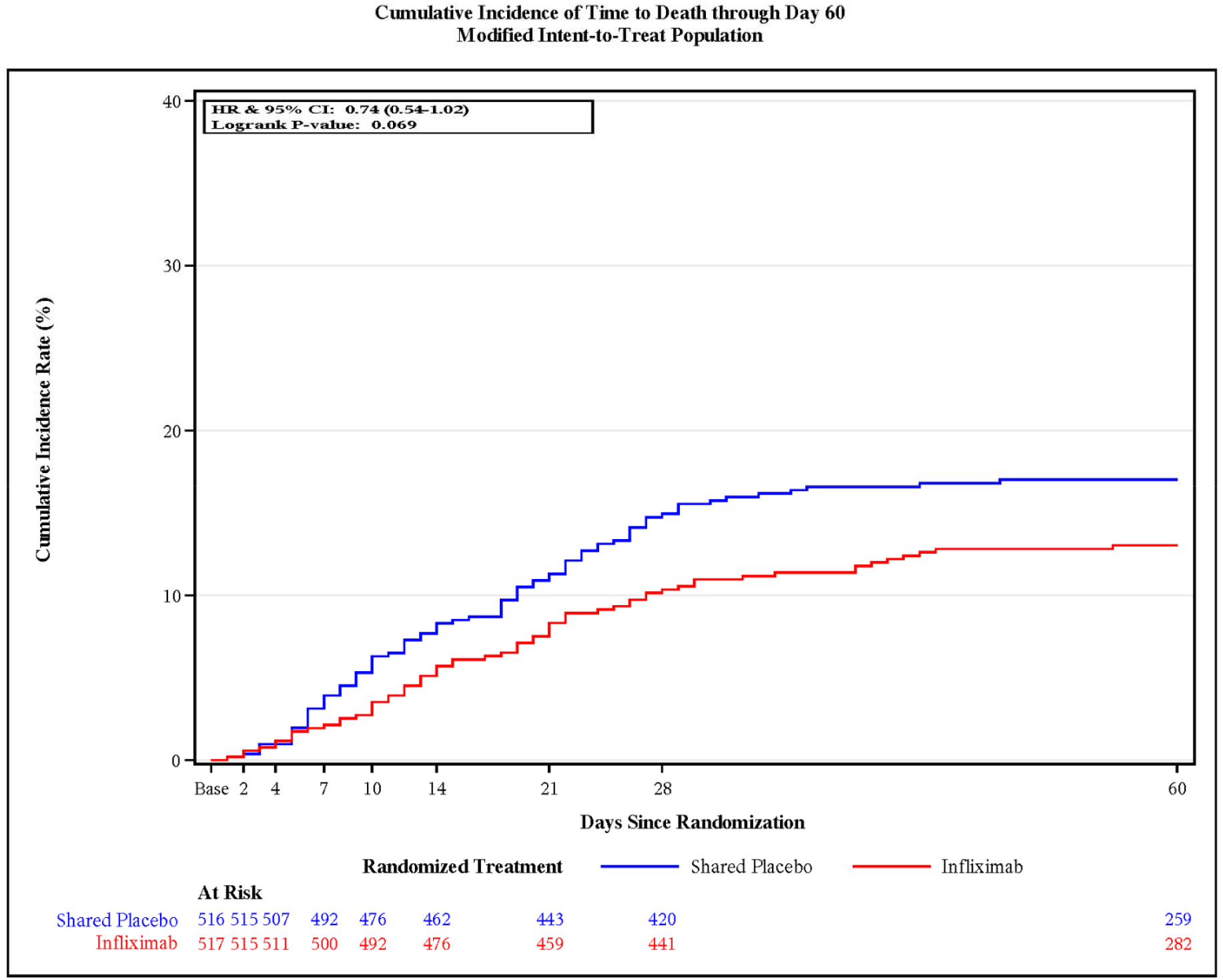
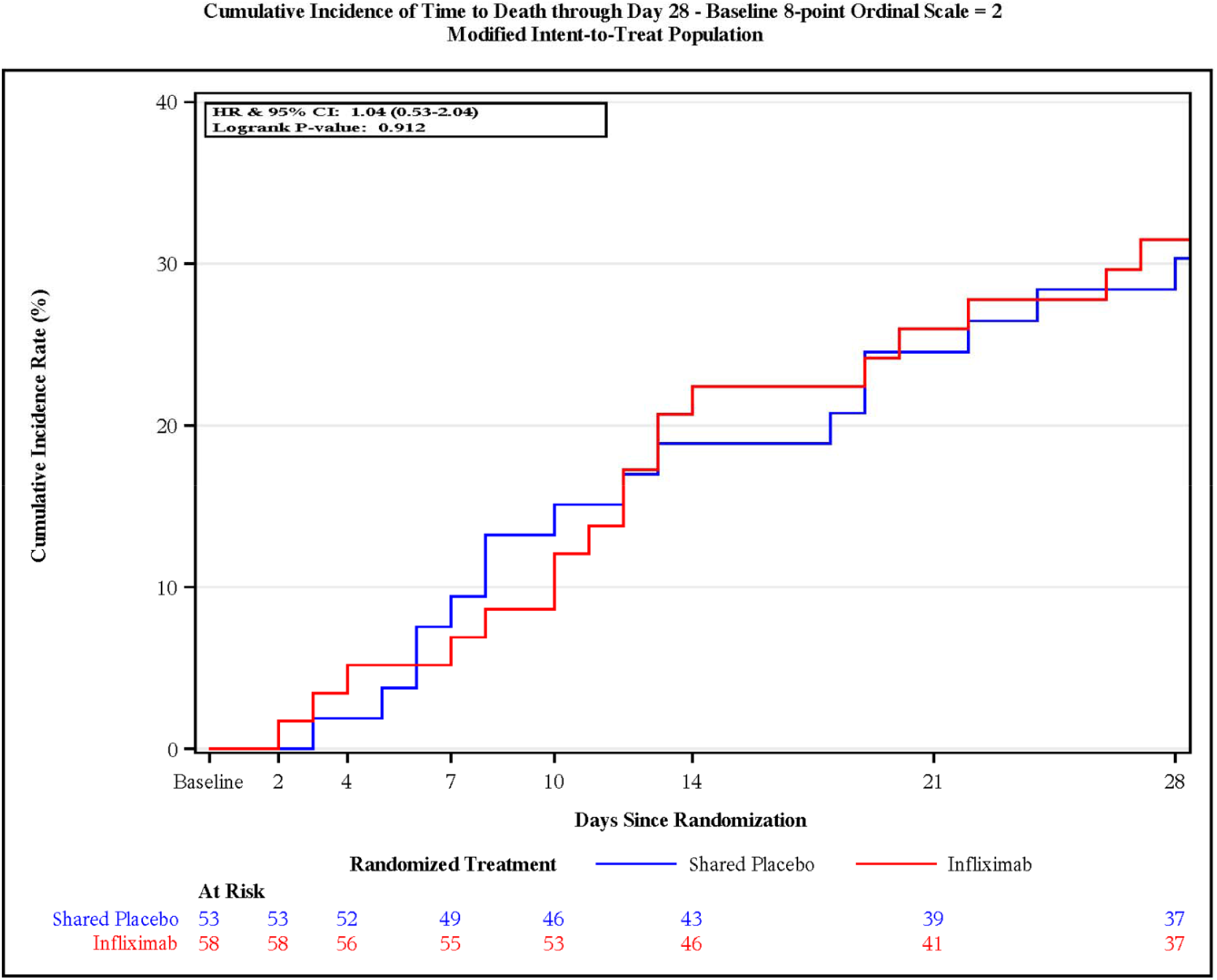
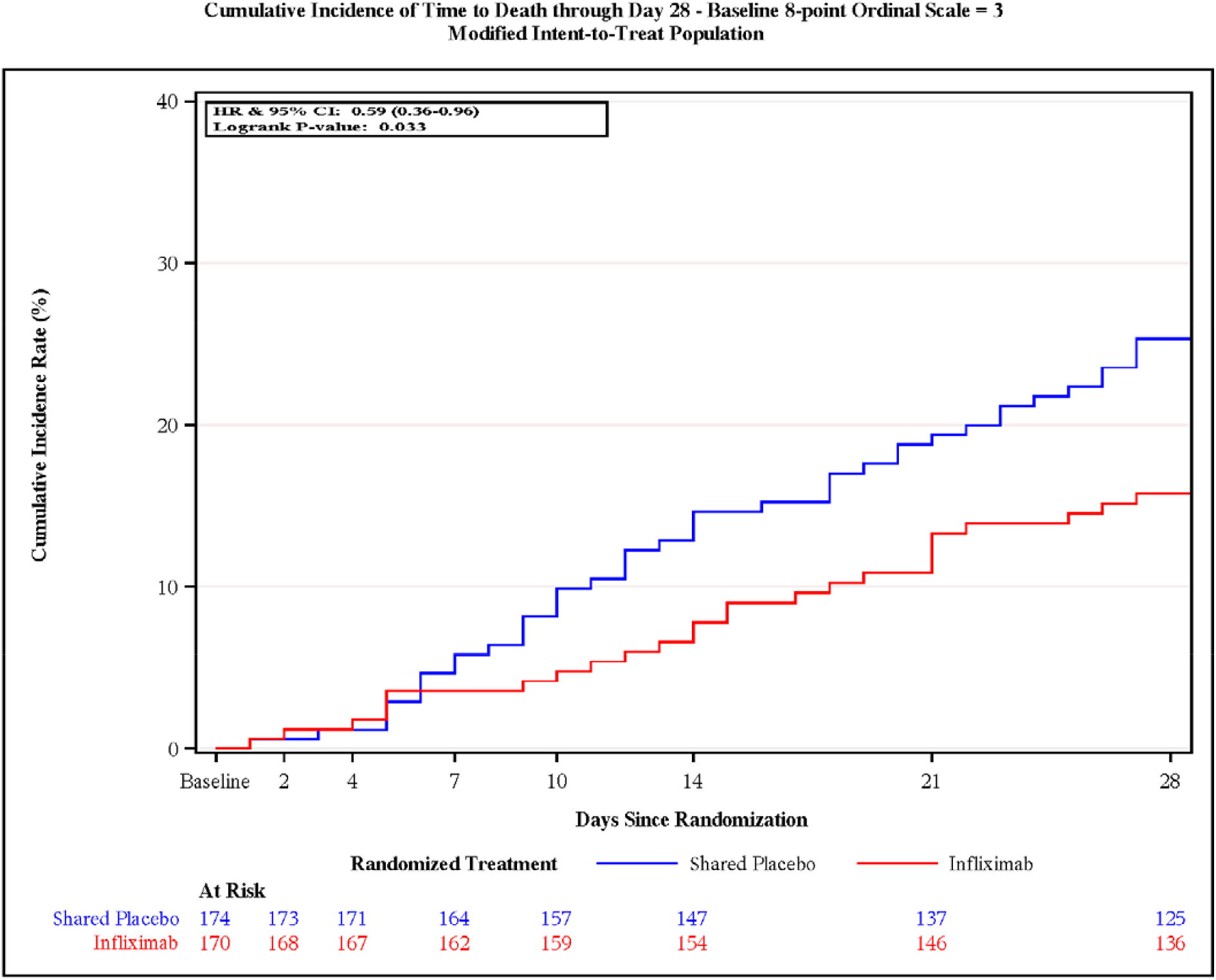
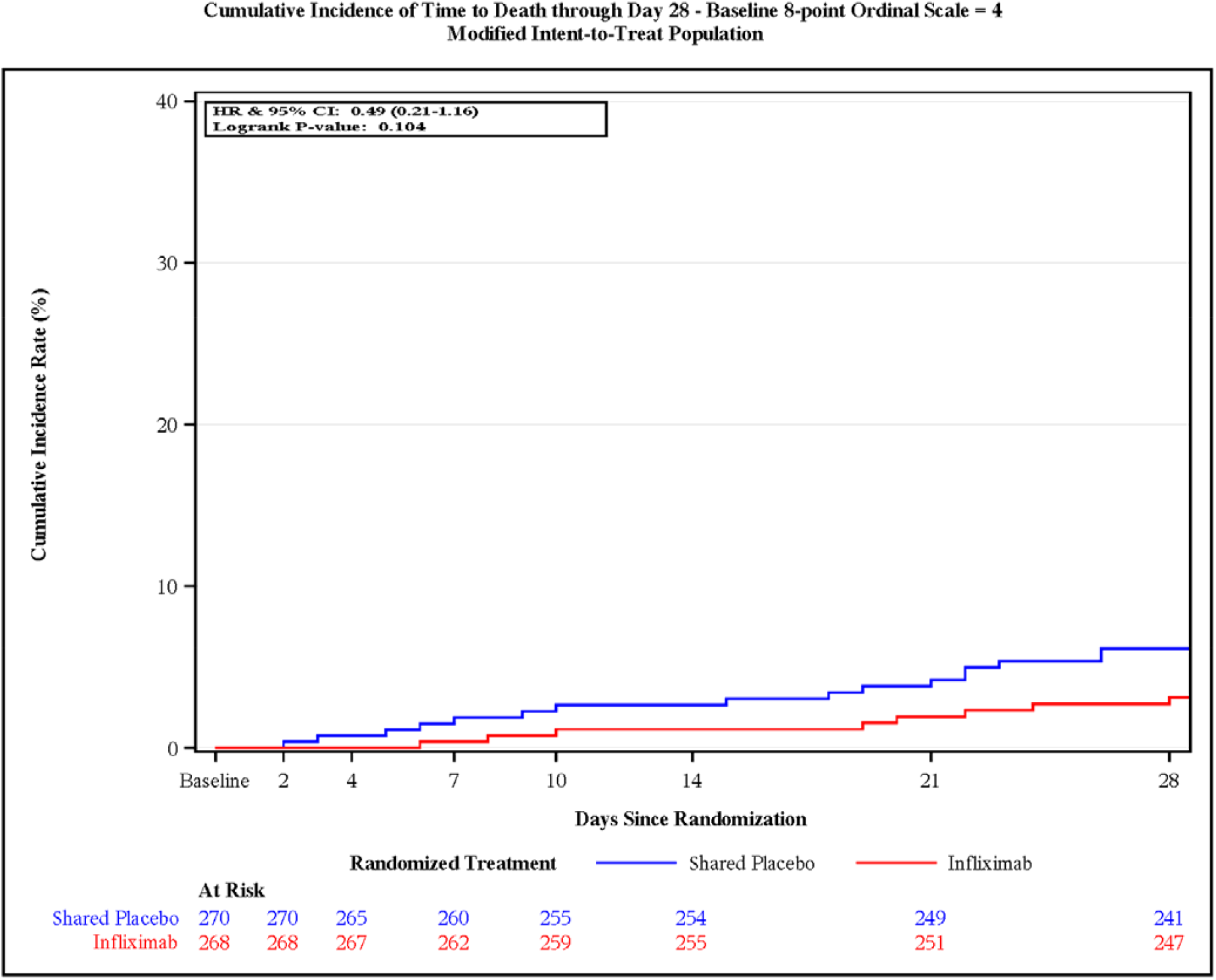
Kaplan-Meier curves of cumulative incidence of overall mortality at (A) day 28, (B) day 60, and by illness severity by ordinal scale for (C) mechanical ventilation or ECMO, (D) high- flow oxygen devices or non-invasive ventilation, and (E) low-flow supplemental oxygen.

#### Clinical status

The odds of improvement in clinical status at day 14 and day 28, as assessed by the OS, were greater with infliximab compared with shared placebo (OR for improvement 1.32, 95% CI 1.05–1.66) and (1.45 95% CI 1.14–1.85) **Tables 2 & S4, Figure S3**. For day 14, the interaction p-value for subgroup analysis by OS was 0.90. On day 14, clinical status was improved with infliximab compared with shared placebo for those at OS 2, 3, and 4 at randomization with the strongest effect in OS 3 (OR 1.48, 95% CI 1.01–2.17). No difference was observed for OS 5. All intention-to-treat data are presented in **Tables S5 and S6**.

### Safety Assessments

No difference was observed in the composite safety endpoint at day 60 (32.9% vs. 33.7%; hazard ratio [HR] 0.96, 95% CI 0.78–1.19). The number of participants with one or more grade 3 or 4 AEs was similar, (infliximab 146 [28.2%], shared placebo 131 [25.4%], risk difference 2.9; 95% CI -2.6 to 8.3) (**Table 3**). SAEs occurred in 125 (24.2%) participants in the infliximab group, with seven events in six participants (1.2%), assessed by investigators as infliximab-related. SAEs occurred in 130 (25.2%) participants in the control group, and the events were attributed to trial product in seven of these participants (1.4%). One participant experienced grade 1 (mild) infusion reaction in the infliximab group.

**Table 3.**
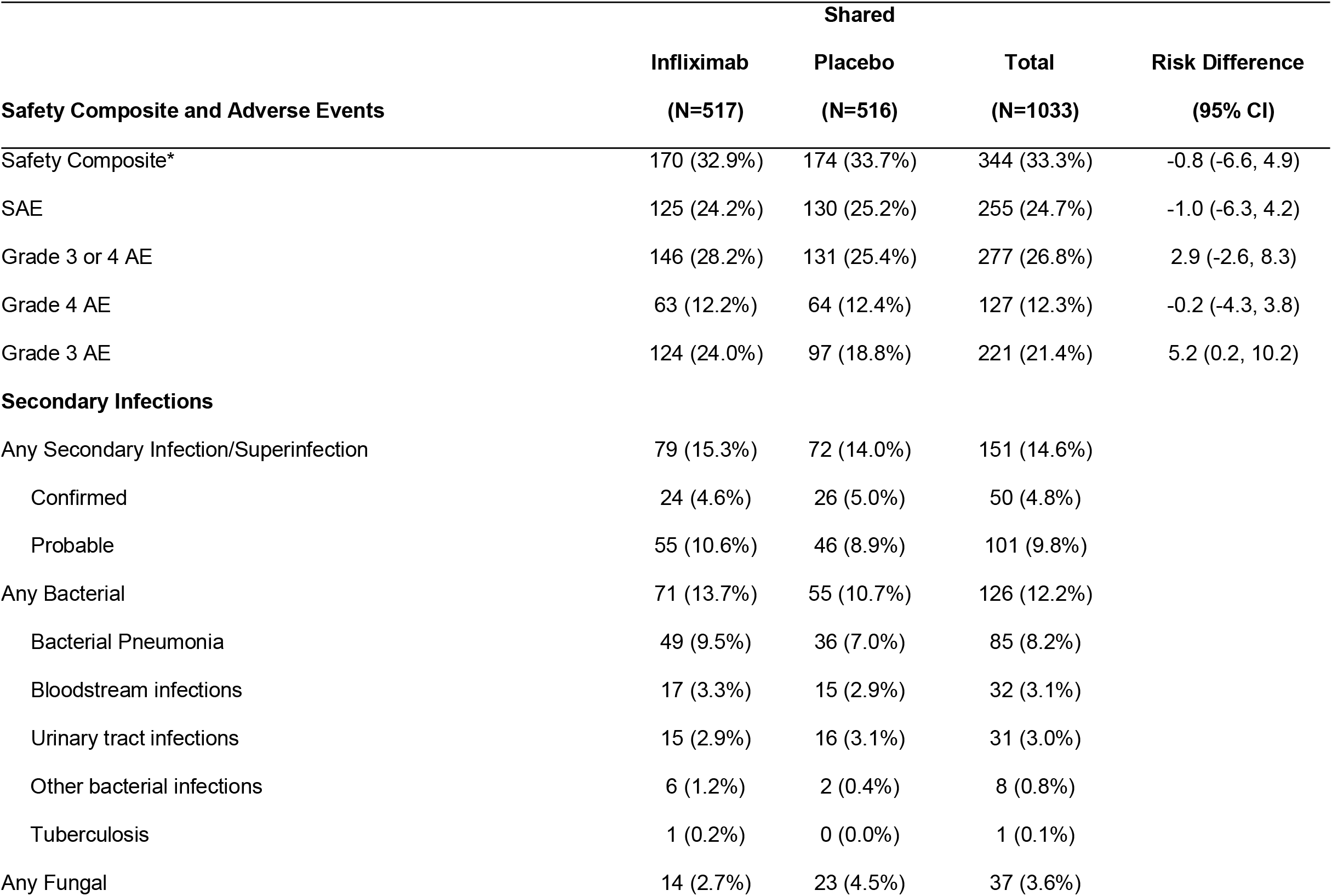
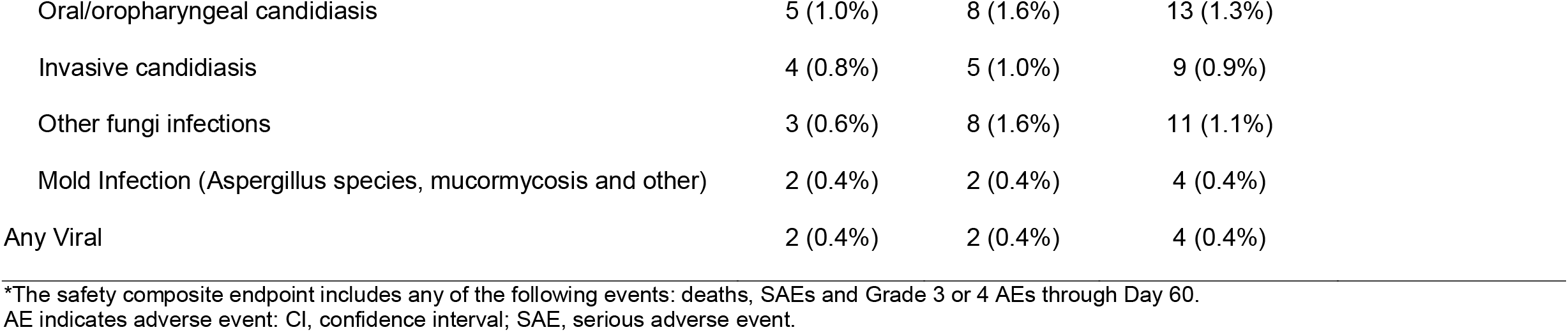
Safety composite and adverse events through day 60.

The percentage of participants who had any secondary infection was similar at day 60 with infliximab and shared placebo (79 [15.3%] vs. 72 [14.0%]) (**Table 3**). The most common secondary infections were bacterial pneumonia, bloodstream and urinary tract infections (**Table 3**). All secondary infections were adjudicated by an independent safety officer.

## DISCUSSION

This randomized, double-blind, placebo-controlled trial evaluated the addition of infliximab to standard of care in hospitalized participants with moderate-to-severe Covid-19 pneumonia. The trial displayed a strong, but not statistically significant, improvement in the primary endpoint of time to recovery. We did, however, observe substantial improvements for key secondary endpoints of 28-day mortality and 14-day clinical status. Infliximab was associated with a 41% lower adjusted odds of death at 28 days in participants hospitalized with Covid-19. The mortality benefit was observed across age groups, sexes, and races/ethnicities. Analysis demonstrated that this mortality benefit was maintained to the completion of study at 60 days. Although some additional deaths occurred after day 28 in both groups, these were mainly in participants intubated at day 28. Sub-analysis by OS showed infliximab, when added to standard of care, reduced mortality across a spectrum of disease severity with hospitalized participants requiring supplemental oxygen and those on high-flow oxygen devices benefiting. In contrast, we observed no benefit in those on mechanical ventilation or ECMO or those not requiring supplemental oxygen. In another subgroup analysis, a stronger mortality benefit was observed with the addition of infliximab to standard of care in participants with CRP >75mg/L at baseline compared with those with CRP ≤75 mg/L.

Identification of anti-inflammatory agents to prevent or reverse dysregulated immune cascades characteristic of severe Covid-19 has are an important area of investigation. The addition of dexamethasone was shown to improve survival in patients requiring supplemental oxygen and became embedded in standard of care.^3^ However, the high morbidity and mortality of Covid-19 and heterogeneity of therapeutic responses suggested the need for additional immunomodulators. The JAK1/JAK2 inhibitor, baricitinib, and IL-6 antibody, tocilizumab, have shown benefit for patients particularly in the setting of progressive respiratory failure.^4,6,16,17^ Results from these trials prompted the NIH guidelines panel to recommend tocilizumab or baricitinib as a second immunomodulator in addition to dexamethasone for patients with progressive respiratory failure and evidence of systemic inflammation.^18^

The infliximab data reported here, and a parallel report of the study of abatacept from ACTIV-1 IM, adds to our knowledge by demonstrating mortality benefit for two separate immunomodulators with unique and different mechanisms of action. Taken together with prior studies, we now have substantial evidence that additional immunomodulation added to corticosteroids reduces mortality and improves clinical outcomes in hospitalized patients with moderate/severe Covid-19. The fact that this occurs with a variety of immunomodulatory agents, all with different targets, is particularly interesting and establishing a better understanding of this synergy is warranted at a mechanistic level.

Infliximab, in particular, and TNF blockade, in general, may be beneficial in Covid-19 management where elevated cytokine levels are seen in severe disease. TNF activates a wide variety of immune cells and aberrant TNF-signaling is a central feature of cytokine release syndrome.^19^ One concern with TNF blockade in the setting of any viral illness is secondary infection. Encouragingly, there were no new or negative safety signals observed during this trial, including no differences between the infliximab and placebo groups in secondary infections.

These observations are consistent with infliximab’s known safety profile having been used in the treatment of inflammatory diseases for over twenty years. Likewise, the simplicity of a single infusion and the global availability of infliximab could potentially increase the arsenal of therapies available for treatment of moderate/severe Covid-19 in settings where current guideline recommended therapies are not widely available.

With each completed trial we gain greater understanding of Covid-19, although numerous clinical questions remain. In particular, there is lack of clarity around the optimal management of patients requiring low-flow oxygen when first hospitalized. Subgroup analyses presented here begin to address this. We show an apparent mortality benefit for immunomodulators in both moderate and severe illness. Our results for participants with moderate disease show improvement independent of inflammatory markers or clinical factors, suggesting treatment with infliximab early in the disease process could provide benefit. In contrast, our data do not support the addition of infliximab to dexamethasone in patients already requiring mechanical ventilation. Additionally, subgroup analysis suggested a greater benefit in both time to recovery and mortality in participants with CRP >75mg/L, suggesting patients with evidence of more severe systemic inflammation at presentation to hospital may benefit more from the addition of a second agent, such as infliximab, to dexamethasone. Future studies should examine if a biomarker-driven approach facilitates early identification of people at-risk for progression who could benefit from additional immunomodulation.

A limitation of this study is that the primary endpoint did not reach statistical significance. Therefore, based on the pre-defined gatekeeping approach, the key secondary endpoints of 28- day mortality and day 14 clinical status are not considered statistically significant although very clinically relevant. A major challenge for this and other studies during the pandemic was the appropriate selection of a primary endpoint in the setting of rapidly changing clinical scenarios.^20^ While a mortality primary endpoint is sometimes considered definitive and lacking in subjectivity, it can require large participant numbers and potentially result in prolonged study recruitment and delayed results. In addition, mortality does not encompass other patient-centered outcomes which are a greater focus in endpoints such as time to recovery or clinical status. These issues represent some factors that led to the selection of the primary endpoint in this study. Secondly, this study was performed in the pre-omicron era. However, despite changes in the predominant circulating variant over time and decreased disease severity overall, we continue to see patients admitted to hospital with respiratory failure and evidence of immune dysregulation. Finally, while there is a theoretical concern that a shared control group could negatively impact multiple treatment evaluations in a platform study such as this, an examination of baseline characteristics does not indicate this issue for ACTIV-1 IM. Integrity of the comparative analyses was preserved through inclusion of only those participants in the shared placebo group who were eligible to receive infliximab, and through the requirement that placebo participants were shared only among agents active in the master protocol at the same time. More importantly, the use of a shared control group has significant practical and ethical benefits.^21^

This report from ACTIV-1 IM shows that infliximab added to standard of care was associated with clinically meaningful improvements in a number of key secondary outcomes, including a 4% absolute reduction in 28-day mortality. As SARS-CoV-2 moves from being pandemic to endemic, and while new variants continue to emerge, ongoing morbidity and mortality from this disease is likely. Expanding our treatment toolbox and developing optimized treatment strategies remains paramount.

## Data Availability

It is expected that the partial de-identified patient dataset will be made available via the 
ACTT Trial Database at NIAID - Access Clinical Data @ NIAID beginning 6 months after publication of the primary results and for 5 years following article publication.

## Acknowledgements

We thank the members of the ACTIV-1 Study Team (see Supplementary Appendix) for their many contributions in conducting the trial, the members of the data and safety monitoring board, William Blackwelder, PhD (chair) (University of Maryland School of Medicine, Baltimore,MD), Timothy G. Buchman, PhD, MD (Emory University School of Medicine, Atlanta, GA), Wilbur H. Chen, MD, MS (University of Maryland School of Medicine, Center for Vaccine Development and Global Health, Baltimore, MD), Lawrence H. Moulton, PhD (Johns Hopkins University, Bloomberg School of Public Health, Baltimore, MD), David M. Parenti, MD (The George Washington University School of Medicine, Washington, DC), Carol O. Tacket, MD (University of Maryland School of Medicine, Center for Vaccine Development and Global Health, Baltimore, MD), William Checkley, MD, PhD (Johns Hopkins University, Baltimore, MD), and Beatriz Grinsztejn, MD (Instituto Nacional de Infectologia Evandro Chagas-Fiocruz, Rio de Janeiro, Brazil) for their oversight; the Community Advisory Board, Larisa Caicedo, Ashish Cowlagi, Anna Davis, Lincoln Larmond, Doug Lindsey, Bob Pearson and the patients and their families for their altruism in participating in this trial.

## Data Sharing

A data sharing statement provided by the authors is available with the full text of this article at NEJM.org.

