## Supplementary Appendix for "Infliximab for Treatment of Adults Hospitalized with Moderate or Severe Covid-19"

The following study group members were all closely involved with the design, implementation, and oversight of the ACTIV-1 study.

**United States Sites**

Jamaica Hospital Medical Center, Jamaica, NY. Mahendra Patel, M.D.

Virginia Commonwealth University Medical Center, Richmond, VA. Arun Sanyal, M.D.

University of Tennessee Medical Center, Knoxville, TN. Jason Green, D.O.

University of Massachusetts Medical Center, Worcester, MA. Jennifer P. Wang, M.D.

University of Oklahoma Health Sciences Center, Oklahoma City, OK. Huimin Wu, M.D., M.P.H.,

Boston Medical Center, Boston, MA. Benjamin Linas, M.D., M.P.H.

Stanford University Medical Center, Palo Alto, CA. Philip Grant, M.D.

Mayo Clinic, Rochester, MN. Vivek Iyer, M.D.

UCLA Medical Center, Los Angeles, CA. Otto Yang, M.D.

Hackensack University Medical Center, Hackensack, NJ. Bindu Balani, M.D.

New York University Langone Medical Center, New York, NY. Sam Parnia, M.D.

University of Arkansas Medical Sciences, Little Rock, AR. Ryan Dare, M.D., M.S.

Wake Forest University Health Sciences, Winston-Salem, NC. Caryn G. Morse, M.D., M.P.H.

University of Utah, Salt Lake City, UT. Estelle S. Harris, M.D.

MedStar Washington Hospital Center, Washington, DC. Glenn Wortmann, M.D.

Tufts Medical Center, Boston, MA. Nicholas Hill, M.D.

UF Health Research, Jacksonville, FL. Shama Patel, M.D.

Ochsner Medical Center, New Orleans, LA. Julia Garcia-Diaz, M.D.

Riverside University Health System, Moreno Valley, CA. Suman Thapamager, M.D.

University of Texas Health Science Center- Tyler, Tyler, TX. Megan Devine, M.D.

Tulane University Health Science Center, New Orleans, LA. Christine M. Bojanowski, M.D., M.S.C.R.

Anne Arundel Medical Center, Annapolis, MD. Barry Meisenberg, M.D.

University of Mississippi Medical Center, Jackson, MS. Gailen Marshall, M.D.

University of Missouri Health System, Columbia, MO. Dima Dandachi, M.D.

Gundersen Health System, La Crosse, WI. Arick Sabin, D.O.

Avera McKennan Hospital & University Health Center, Sioux Falls, SD. Anthony Breemo, M.D.

Trinity Mother Frances Hospital, Tyler TX. Suman Sinha, M.D.

University of Washington Medical Center, Seattle, WA. Christopher Goss, M.D.

West Virginia University, Morgantown, WV. Rebecca Reece, M.D.

Mercy St. Vincent Medical Center, Toledo, OH. Arlette Aouad, M.D.

University of Buffalo, Buffalo, NY. Seth Glassman, M.D.

University of Kentucky, Lexington, KY. Peter Morris, M.D.

The University of Texas Health Science Center – Houston, Houston, TX. Bela Patel, M.D.

University of Texas - Rio Grande Valley, Weslaco, TX. Fatimah Bello, M.D.

**Ex-US sites**

Hospital Ernesto Dornelles, Porto Alegre, Brazil. Juliana Cardozo Fernandes, M.D.,

Hospital de Chancay, Lima, Peru. Oscar Carbajal, M.D.

Hospital Rawson, Cordoba, Argentina. Lorena Ravera M.D.

Hospital Felcio Rocho, Belo Horizonte, Brazil. Mozar Castro M.D.

Hospital Regional Lambayaque, Chiclayo, Peru. Miguel Villegas-Chiroque, M.D.

Sanatorio Allende, Cordoba, Argentina. Fernando Oscar Riera, M.D.

Hospital Universitario Dr. Jose Eleuterio Gonzalez, Nuevo Leon, Mexico. Adrian Camacho, M.D.

Santa Casa de Misericordia de Porto Alegre, Porto Alegre, Brazil. Claudio Stadnik M.D.

Hospital Nacional Arzobispo Loayza, Lima, Peru. Jorge Gave, M.D.

Hospital Brasilia, Brasilia, Brazil. Rodrigo Biondi. M.D.

Hospital Santa Rosa, Piura, Peru. Ronal Gamarra Velarde, M.D.

Instituto D'OR de Ensino e Pesquisa, Rio de Janeiro, Brazil. Jose Cerbino Neto, M.D.

Hospital Interzonal Dr Jose Penna Bahia Blanca, Bahia Blanca, Argentina. Juan Ditondo, M.D.

Hospital Ramos Mejia, Buenos Aires, Argentina. Marcelo H. Losso, M.D.

Sanatorio Ramon Cereijo, Caba, Argentina. Mariano Dolz, M.D.

**Other Contributors**

AbbVie, North Chicago, IL. William Ferguson, PhD

Bristol Myers-Squibb, New York, NY. Michael Maldonado, MD

Janssen Pharmaceuticals/Johnson and Johnson, Beerse, Belgium. Maria Beumont-Mauviel, MD

Gilead, Foster City, CA. Huyen Cao, M.D.,

Duke Clinical Research Institute (DCRI), Durham, NC. Rose Beci, Jun Wen, Sean O’Brien, PhD

Technical Resources International, Inc (TRI), Bethesda, MD. Daniel Molina, PharmD, MBA; Sandhya Rao, MD, M.Ch.

Health and Human Services, Washington, DC. Thomas Stock, D.O., William Erhardt, M.D. (H-CORE, formally Operation Warp Speed), Sarah Read, M.D., (National Institute of Allergy and Infectious Diseases)

National Center for Advancing Translational Sciences, Bethesda, MD. Jessica Springer, RN, MPH/MPA,

### **ACTIV-1 Study Team Members**

The ACTIV-1 study was a significant collaborative effort across many countries, partners and sites. Below is a list of sites and staff that worked tirelessly to implement and execute the ACTIV-1 Trial.

**United States Sites**

Washington University St. Louis, St Louis, MO. William G. Powderly, M.D., Jane A. O’Halloran, M.D., Rachel Presti, M.D., Ph.D., Adriana Rauseo Acevedo, M.D., Luis Parra Rodriguez, M.D.

Duke University Medical Center, Durham, NC. Cameron Wolfe, M.B.B.S., M.P.H., Emily Ko, M.D., Ph.D., Tatyana Der, M.D., Lana Wahid, M.D.

Weill Cornell Medical College, New York, NY. Matthew W. McCarthy, M.D., Elizabeth Salsgiver, Kate Willsey, Candace Alleyne, Britta Witting.

Robert Wood Johnson Medical School, New Brunswick, NJ. Reynold Panettieri, M.D.

Jamaica Hospital Medical Center, Jamaica, NY. Mahendra Patel, M.D.

Oregon Health and Science University, Portland, OR. Akram Khan, M.D.

University of Illinois at Chicago, Chicago, IL. Alfredo J. Mena Lora, M.D., Rodrigo Burgos, Pharm.D., Brenna Lindsey, M.P.H., Fischer Herald, Pharm.D.

University of Kansas Medical Center, Kansas City, KS. Matthias Salathe, M.D., Charles D. Bengtson, M.D., Andreas Schmid, M.D., Kimberly Lovell, Ph.D.

St. Lawrence Health System, Potsdam, NY. Eyal Kedar, M.D., Carly Lovelett, C.C.R.P., M.S., M.B.A., Daniel Soule, D.O., Daniel Jaremczuk, C.C.R.P.

Trinitas Hospital, Elizabeth, NY. Gerardo Capo, M.D.

University of Texas Health Science Center at San Antonio, San Antonio, TX. Thomas F. Patterson, M.D., Heta Javeri, M.D., Philip O. Ponce, M.D., Danielle O. Dixon, D.O.

University of Rochester Medical Center-Strong Memorial Hospital, Rochester, NY. Christopher Palma, M.D., Sc.M., Caroline M. Quill, M.D.

University of North Carolina Hospital, Chapel Hill, NC. Anne Lachiewicz, M.D.

Virginia Commonwealth University Medical Center, Richmond, VA. Arun Sanyal, M.D.

MidMichigan Medical Center – Midland, Midland, MI. John Blamoun, M.D.

University of Tennessee Medical Center, Knoxville, TN. Jason Green, D.O., Abby Smith, R.N., Glenda Brown, R.N.

Providence Medical Research Center, Spokane, WA. Radica Alicic, M.Sc., M.D., Kyle Varner, M.D., Joni Baxter, R.N., Tracy Roundy, B.S.

University of Massachusetts Medical Center, Worcester, MA. Jennifer P. Wang, M.D., Mary Co, M.D., Mireya Wessolossky, M.D., Juan Perez-Velazquez, M.D.

University of Oklahoma Health Sciences Center, Oklahoma City, OK. Huimin Wu, M.D., M.P.H., Jennifer Holter-Chakrabarty, M.D., Brittany Karfonta, Juvaria Anjum.

Boston Medical Center, Boston, MA. Benjamin Linas, M.D., M.P.H., Jai Marathe, M.D., Myriam Castagne, Daniel MomPoint.

Stanford University Medical Center, Palo Alto, CA. Philip Grant, M.D.

Mayo Clinic, Rochester, MN. Vivek Iyer, M.D.

UCLA Medical Center, Los Angeles, CA. Otto Yang, M.D.

Hackensack University Medical Center, Hackensack, NJ. Bindu Balani, M.D., Marylynn Breslin.

New York University Langone Medical Center, New York, NY. Sam Parnia, M.D.

University of Arkansas Medical Sciences, Little Rock, AR. Ryan Dare, M.D., M.S., Susan Dodson, M.B.A., B.S.N., R.N.

Wake Forest University Health Sciences, Winston-Salem, NC. Caryn G. Morse, M.D., M.P.H., John Williamson, Pharm.D.

University of Utah, Salt Lake City, UT. Estelle S. Harris, M.D., Elizabeth A. Middleton, M.D. MedStar Washington Hospital Center, Washington, DC. Glenn Wortmann, M.D.

Tufts Medical Center, Boston, MA. Nicholas Hill, M.D.

UF Health Research, Jacksonville, FL. Shama Patel, M.D.

Ochsner Medical Center, New Orleans, LA. Julia Garcia-Diaz, M.D.

Riverside University Health System, Moreno Valley, CA. Suman Thapamager, M.D.

University of Texas Health Science Center- Tyler, Tyler, TX. Megan Devine, M.D.

Tulane University Health Science Center, New Orleans, LA. Christine M. Bojanowski, M.D., M.S.C.R.

Anne Arundel Medical Center, Annapolis, MD. Mai Tavadze, M.D., Barry Meisenberg, M.D.

University of Mississippi Medical Center, Jackson, MS. Gailen Marshall, M.D.

University of Missouri Health System, Columbia, MO. Dima Dandachi, M.D.

Gundersen Health System, La Crosse, WI. Arick Sabin, D.O.

Avera McKennan Hospital & University Health Center, Sioux Falls, SD. Anthony Breemo, M.D., Nate Miller, M.D.

Trinity Mother Frances Hospital, Tyler TX. Suman Sinha, M.D.

University of Washington Medical Center, Seattle, WA. Christopher Goss, M.D.

West Virginia University, Morgantown, WV. Rebecca Reece, M.D.

Mercy St. Vincent Medical Center, Toledo, OH. Arlette Aouad, M.D.

University of Buffalo, Buffalo, NY. Seth Glassman, M.D.

University of Kentucky, Lexington, KY. Peter Morris, M.D.

The University of Texas Health Science Center – Houston, Houston, TX. Bela Patel, M.D., Pratik Doshi, M.D.

University of Texas - Rio Grande Valley, Weslaco, TX. Fatimah Bello, M.D.

**Ex-US sites**

Hospital Nacional Hipolito Unanue, Lima, Peru. Patricia Segura Nuñez,M.D., Pablo Torres, M.D., Ricardo Sanchez, M.D., Gladys Murga, M.D.

Hospital Central de la Fuerza Aerea del Peru, Lima, Peru. Sabina Mendiviltuchia de Tai, M.D., Johana Calderon Galvez, M.D., Maria Lyda Icochea Perez, M.D., Claudia Carolina Becerra Nunez, M.D.

Hospital Ernesto Dornelles, Porto Alegre, Brazil. Juliana Cardozo Fernandes,M.D., M.Sc., Felipe Ceriolli Breda, M.D., Mauricio Mello Roux Leite, M.Sc., Tobias Milbradt, M.Sc.

Sanatorio Diagnostico, Santa Fe, Argentina. Martin Maillo, M.D., Luz Rodeles, M.D., Ph.D., Nadia Benzaquen, M.D., Sebastian Pezzini, M.D.

Nuevo Hospital Civil de Guadalajara Juan I. Menchaca, Guadalajara, Mexico. Daniel Rodríguez Gonzalez, M.D., Iliana Higareda Almaraz, M.D., Víctor Hugo Madrigal Robles, M.D., María Fernanda Rosas Ismerio, M.D.

Clinica Central S.A., Villa Regina, Argentina. Horacio Ariza, M.D., Juan Manuel Calderon, M.D., Eduardo Borsetta, M.D., Noemí Sandoval, B.Q.

Hospital e Maternidade Celso Pierro - PUC Campinas, Campinas, Brazil. Maria Patelli Juliani Souza Lima, M.D.

Hospital de Clinicas de Porto Alegre HCPA, Porto Alegre, Brazil. Eduardo Sprinz, M.D., Laura De Bona,B.Q., Maria Eduarda Claus, Nut., Arthur Pille, M.D.

Sanatorio ritánico, Rosario, Argentina. Matías Lahitte, M.D., Mariángeles Fenés M.D., Cecilia Bianchi M.D., María Emilia Miserere M.D.

Instituto Medico Platense, La Plata, Argentina. Analia Mykietiuk, M.D.

Hospital de Chancay, Lima, Peru. Oscar Carbajal, M.D.

Hospital Rawson, Cordoba, Argentina. Lorena Ravera M.D.

Hospital Felcio Rocho, Belo Horizonte, Brazil. Mozar Castro M.D.

Hospital Regional Lambayaque, Chiclayo, Peru. Miguel Villegas-Chiroque, M.D., Halbert Christian Sanchez Carrillo, M.D.

Sanatorio Allende, Cordoba, Argentina. Fernando Oscar Riera, M.D., Aldana Mano, M.D.

Hospital Universitario Dr. Jose Eleuterio Gonzalez, Nuevo Leon, Mexico. Adrian Camacho, M.D.

Santa Casa de Misericordia de Porto Alegre, Porto Alegre, Brazil. Claudio Stadnik M.D.

Hospital Nacional Arzobispo Loayza, Lima, Peru. Jorge Gave, M.D.

Hospital Brasilia, Brasilia, Brazil. Rodrigo Biondi. M.D.

Hospital Santa Rosa, Piura, Peru. Ronal Gamarra Velarde, M.D.

Instituto D'OR de Ensino e Pesquisa, Rio de Janeiro, Brazil. Jose Cerbino Neto, M.D.

Hospital Interzonal Dr Jose Penna Bahia Blanca, Bahia Blanca, Argentina. Juan Ditondo, M.D., Myrna Zuain, M.D.

Hospital Ramos Mejia, Buenos Aires, Argentina. Marcelo H. Losso, M.D., Javier J. Toibaro, M.D.

Sanatorio Ramon Cereijo, Caba, Argentina. Mariano Dolz, M.D.

**Other Contributors**

Companies that helped with protocol in the earlier phase of development as part of the ACTIV working group.

AbbVie, North Chicago, IL. David B. Bharucha, M.D., Ph.D., FACC, William Ferguson, Ph.D.

Bristol Myers-Squibb, New York, NY. Brian Gavin, Ph.D., Jonathan Sadeh, M.D. MSc, Sheila Kelly, MD, Michael Maldonado, M.D.

Janssen Pharmaceuticals/Johnson and Johnson, Beerse, Belgium. Rich Melsheimer, Maria Beumont-Mauviel, M.D., Marita Stevens, M.D.

Gilead, Foster City, CA. Huyen Cao, MD, Adam DeZure, MD, Kavita Juneja, MD, Mazin Abdelghany, MD

Duke Clinical Research Institute (DCRI), Durham, NC. Daniel Benjamin, MD, MPH, PhD, Brian Smith, MD MPH MHS, Theresa Jasion, Rachel Olson, RN MS MBA, Megan Roebuck, M.S., Jacqueline Huvane, Ph.D., Steve McNulty, Pearl Zakroysky, Jun Wen, Adam Silverstein, Jeff Leimberger, Eric Yow, Zhen Huang, Hwasoon Kim, Carla Anderson, Carrie Elliott RN, Merri Swartz RN, Rose Beci, Susan Halabi, Jyotsna Garg, Hussein Al-Khalidi, Sean O’Brien, PhD

Technical Resources International, Inc (TRI), Bethesda, MD. Sandra Butler, Ph.D., Daniel Molina, PharmD, MBA; Neta Nelson, MPH, Divya Kalaria, MsCRA, MsFM, Sandhya Rao, MD, M.Ch, Ketty Philogene, MD, Tim Schulz, Ph.D; Averie Kuek, Fatou Bah, MSHS; Jarrard Mitchell, MBA; Elizabeth Polo, M.D., Michelle Wong, J.D.

Syneos Health, Morrisville, NC. Sharon Baldan, Sandra Mendez, MD, Bradford Stevens, Marcela Toledo, Talita Abba, Emma Herrejon, R.Ph; Cristina Gomez

Fisher Clinical Services, Rockville, MD. Georgeta Mardari, MS, MBA; Neeraja Putta, R.Ph, MBA

University of North Carolina, Chapel Hill, NC Kevin Anstrom, Ph.D. Lisa LaVange, Ph.D.

Health and Human Services, Washington, DC Thomas Stock, D.O., Williaml Erhardt, M.D. (H-CORE, formally Operation Warp Speed), Sarah Read, M.D., Mike Proschan, Ph.D. (National Institute of Allergy and Infectious Diseases)

Foundation of the National Institutes of Health, North Bethesda, MD. Stacey Adam, Ph.D.

Biomedical Advanced Research and Development Authority, Washington, D.C. Robin Mason, MS, MBA, Holli Hamilton, M.D.; Derek Eisnor, M.D., Anna O’Rourke, MS, Aditi Patel, Ph.D., RAC, Betty Brody, Anna Chiang, MSW, MPA,

National Center for Advancing Translational Sciences, Bethesda, M.D. Sam Bozzette, M.D., Ph.D., Soju Chang, M.D., MPH, Cynthia Boucher, MS, Jessica Springer, RN, MPH/MPA, Jane Atkinson, D.D.S., Brian Lind, Lilli M. Portilla, MPA, Ami D. Gadhia, JD, LLM, CLP, Sury Vepa, Ph.D., JD, Emily Carlson Marti, MA, Bobbi Gardner, Joni Rutter, Ph.D., Clare Schmitt, Ph.D., Michael Kurilla, M.D.

### **Supplemental Methods**

**Eligibility**

Participants were included if they were adults ≥18 years of age, with confirmed moderate/severe SARS-CoV 2 infection admitted to hospital or awaiting admission in the emergency department. Participants were required to have radiological evidence of pulmonary involvement or oxygen saturation ≤94% on room air or requirement for supplemental oxygen, mechanical ventilation or extracorporeal membrane oxygenation (ECMO). Participants were excluded if they had elevated liver function tests (AST or ALT >10 times the upper limit of normal), acute kidney injury (estimated glomerular filtration rate <30 ml/min; stable chronic renal insufficiency with eGFR <30 ml/min was permitted), history of New York Heart Association class III/IV congestive heart failure, neutropenia (<1.0 x 10^3^/µl), lymphopenia (<0.2 x 10^3^/µl), receipt of targeted immune therapies for any indication in the last four weeks or five drug half-lives including IL-6 and JAK inhibitors (corticosteroids, monoclonal antibodies or convalescent plasma for COVID-19 treatment were permitted), or evidence of untreated tuberculosis or other untreated infections.

**Study oversite**

Institutional review boards at each site or a centralized institutional review board, where applicable, approved the trial protocol. The trial was overseen by an independent data and safety monitoring board (DSMB). Three interim analyses were planned to allow the DSMB to (i) assess the futility of each agent, and (ii) to assess early stopping for efficacy but only two were conducted due to rapid enrollment. Written informed consent was obtained from each participant or their legally authorized representative.

**Randomization**

Firstly, each participant was randomized with equal probability to one of the agents available at the time of enrollment after applying agent-specific safety exclusions. This randomization was open-label. Secondly, each participant was assigned in a blinded manner to the test agent or its matching placebo in an n:1 ratio, where n = the number of agents that the participant was eligible for. Randomization was stratified by geographic location (four United States regions and four Latin American countries) and disease severity (moderate and severe). Moderate disease was defined as hospitalized with or without supplemental oxygen requirement; severe disease was defined as hospitalized requiring high flow oxygen, non-invasive or mechanical ventilation or ECMO.

**Remdesivir administration**

Participants receiving remdesivir at enrollment continued to receive treatment through study supply. Those not on remdesivir at enrollment were initiated as part of study protocol where eligible. Remdesivir was given as an IV loading dose of 200mg followed by a maintenance dose of 100mg up to 10 days.

**Description of the placebo group**

The placebo group for each sub-study only included participants that were eligible for enrollment in that sub-study. If a participant was eligible for all three sub-studies, then that participant was included in the placebo group for all three studies. However, if for example, the participant did not meet agent specific criteria for infliximab, they were not included in the placebo for the infliximab sub-study but could be included in the placebo group for other agents. As a result, the placebo groups for each sub-study were similar but not identical.

**Statistical analysis**

***Sample size***

The sample size requirements were based on the ability to detect a moderate improvement in time to recovery. A total of 788 recoveries were required to provide approximately 85% power to detect a recovery rate ratio (RRR) of 1.25, assuming 73% of participants achieve recovery on standard of care plus placebo, consistent with the ACTT-1 results.^1^ RRR represents the ratio of infliximab to placebo instantaneous rates of recovery.

***Safety assessment***

Related SAEs were reviewed in a timely manner by the study safety review team and DSMB

***Subgroup analyses***

Pre-specified subgroup analyses evaluated treatment effects across geographic region, symptom duration prior to enrollment, baseline disease severity (stratified by moderate versus severe disease, and by ordinal scale category) age, race, sex, C-reactive protein (CRP) and comorbidities. P-values for interaction were calculated for subgroup analyses to identify if the effect of the intervention on the outcome differed between the subgroups.

### **Table S1. P-value cut offs, infliximab versus shared placebo**

| **Endpoint** | **Efficacy boundary -**  **t-statistic and p-value** | **Observed data -**  **t-statistic and nominal p-value** | **Decision based on the gatekeeper approach** |
| --- | --- | --- | --- |
| Time-to-recovery | t>1.9873  p<0.0469 | t=1.8582  p=0.0631 | Fail to reject the null hypothesis and stop the hierarchical testing process |
| 28-day mortality endpoint | t<-2.5624  p<0.0104 | t=-2.4328  p=0.0150 |  |
| 14-day 8-point ordinal scale | t>2.2839  p<0.0224 | t=2.3514  p=0.0187 |  |

| **Interim Analysis** | **Information Fraction** |
| --- | --- |
| First Interim Analysis | 0.3135 |
| Second Interim Analysis | 0.6371 |
| Final Analysis | 1.0000 |

### **Table S2. Demographics and baseline characteristics – US (modified intent-to-treat)**

| **Demographics and baseline characteristics** | **Infliximab**    **(N=330)** | **Shared Placebo**    **(N=331)** | **Total**   **(N=661)** |
| --- | --- | --- | --- |
| Age in years* | 56.0 (14.59) | 56.6 (14.38) | 56.3 (14.48) |
| Male sex | 200 (60.6%) | 179 (54.1%) | 379 (57.3%) |
| Race |  |  |  |
| White | 206 (62.4%) | 216 (65.3%) | 422 (63.8%) |
| Black or African American | 68 (20.6%) | 65 (19.6%) | 133 (20.1%) |
| American Indian or Alaska Native | 3 (0.9%) | 4 (1.2%) | 7 (1.1%) |
| Asian | 11 (3.3%) | 16 (4.8%) | 27 (4.1%) |
| Native Hawaiian or Other Pacific Islander | 1 (0.3%) | 0 (0.0%) | 1 (0.2%) |
| Multiracial | 2 (0.6%) | 1 (0.3%) | 3 (0.5%) |
| Other | 13 (3.9%) | 7 (2.1%) | 20 (3.0%) |
| Unknown | 26 (7.9%) | 22 (6.6%) | 48 (7.3%) |
| Hispanic or Latino Ethnicity | 76 (23.0%) | 76 (23.0%) | 152 (23.0%) |
| Disease severity at baseline^†^ |  |  |  |
| Severe disease | 128 (38.8%) | 136 (41.1%) | 264 (39.9%) |
| Moderate disease | 202 (61.2%) | 195 (58.9%) | 397 (60.1%) |
| Clinical status (8-point ordinal scale) at baseline |  |  |  |
| 1. Death | 0 | 0 | 0 |
| 2. Hospitalized, on invasive ventilation or ECMO | 34 (10.3%) | 27 (8.2%) | 61 (9.2%) |
| 3. Hospitalized, on non-invasive ventilation or high flow oxygen devices | 94 (28.5%) | 109 (32.9%) | 203 (30.7%) |
| 4. Hospitalized, requiring supplemental oxygen | 187 (56.7%) | 181 (54.7%) | 368 (55.7%) |
| 5. Hospitalized, not requiring supplement oxygen, requiring ongoing medical care | 15 (4.5%) | 14 (4.2%) | 29 (4.4%) |
| 6. Hospitalized, not requiring supplement oxygen, not requiring ongoing medical care | 0 | 0 | 0 |
| 7. Not hospitalized, limitations on activity and/or requiring home oxygen | 0 | 0 | 0 |
| 8. Not hospitalized, no limitations on activities | 0 | 0 | 0 |
| Geographic Region |  |  |  |
| USA - Northeast | 119 (36.1%) | 118 (35.6%) | 237 (35.9%) |
| USA - Midwest | 81(24.5%) | 77 (23.3%) | 158 (23.9%) |
| USA - South | 93 (28.2%) | 94 (28.4%) | 187 (28.3%) |
| USA - West | 37 (11.2%) | 42 (12.7%) | 79 (12.0%) |
| Days from Symptom Onset* | -9.2 (4.70) | -9.3 (6.29) | -9.2 (5.55) |
| Body Mass Index (kg/m^2^)* | 33.6 (8.88) | 34.3 (9.01) | 33.9 (8.94) |
| **Comorbidities** |  |  |  |
| Hypertension | 157 (47.6%) | 159 (48.0%) | 316 (47.8%) |
| Obesity (BMI>=30 kg/m^2^) | 200 / 320 (62.5%) | 210 / 317 (66.2%) | 410 / 637 (64.4%) |
| Diabetes Mellitus | 107 (32.4%) | 111 (33.5%) | 218 (33.0%) |
| Coronary Artery Disease | 32 (9.7%) | 24 (7.3%) | 56 (8.5%) |
| History of Heart Failure | 14 (4.2%) | 12 (3.6%) | 26 (3.9%) |
| History of Cancer | 30 (9.1%) | 30 (9.1%) | 60 (9.1%) |
| Asthma | 29 (8.8%) | 43 (13.0%) | 72 (10.9%) |
| Chronic Obstructive Pulmonary Disease | 23 (7.0%) | 20 (6.0%) | 43 (6.5%) |
| Tuberculosis | 2 (0.6%) | 2 (0.6%) | 4 (0.6%) |
| HIV/AIDS | 1 (0.3%) | 4 (1.2%) | 5 (0.8%) |
| Severe Liver Disease | 2 (0.6%) | 2 (0.6%) | 4 (0.6%) |
| Severe Kidney Disease | 3 (0.9%) | 6 (1.8%) | 9 (1.4%) |

 * Displayed as Mean (Std. Dev.)

^†^Disease severity at baseline calculated as moderate disease = ordinal 5 + ordinal 4, severe disease = ordinal 3 + ordinal 2

### **Table S3. Demographics and baseline characteristics – Latin America (modified intent-to-treat)**

| **Demographics and baseline characteristics** | **Infliximab**    **(N=187)** | **Shared Placebo**    **(N=185)** | **Total**    **(N=372)** |
| --- | --- | --- | --- |
| Age in years* | 52.5 (15.33) | 52.0 (14.68) | 52.2 (14.99) |
| Male sex | 125 (66.8%) | 119 (64.3%) | 244 (65.6%) |
| Race |  |  |  |
| White | 113 (60.4%) | 117 (63.2%) | 230 (61.8%) |
| Black or African American | 9 (4.8%) | 3 (1.6%) | 12 (3.2%) |
| American Indian or Alaska Native | 2 (1.1%) | 0 (0.0%) | 2 (0.5%) |
| Asian | 0 (0.0%) | 0 (0.0%) | 0 (0.0%) |
| Native Hawaiian or Other Pacific Islander | 0 (0.0%) | 0 (0.0%) | 0 (0.0%) |
| Multiracial | 0 (0.0%) | 1 (0.5%) | 1 (0.3%) |
| Other | 63 (33.7%) | 64 (34.6%) | 127 (34.1%) |
| Unknown | 0 (0.0%) | 0 (0.0%) | 0 (0.0%) |
| Hispanic or Latino Ethnicity | 176 (94.1%) | 174 (94.1%) | 350 (94.1%) |
| Disease severity at baseline^†^ |  |  |  |
| Severe disease | 100 (53.5%) | 91 (49.2%) | 191 (51.3%) |
| Moderate disease | 87 (46.5%) | 94 (50.8%) | 181 (48.7%) |
| Clinical status (8-point ordinal scale) at baseline |  |  |  |
| 1. Death | 0 | 0 | 0 |
| 2. Hospitalized, on invasive ventilation or ECMO | 24 (12.8%) | 26 (14.1%) | 50 (13.4%) |
| 3. Hospitalized, on non-invasive ventilation or high flow oxygen devices | 76 (40.6%) | 65 (35.1%) | 141 (37.9%) |
| 4. Hospitalized, requiring supplemental oxygen | 81 (43.3%) | 89 (48.1%) | 170 (45.7%) |
| 5. Hospitalized, not requiring supplement oxygen, requiring ongoing medical care | 6 (3.2%) | 5 (2.7%) | 11 (3.0%) |
| 6. Hospitalized, not requiring supplement oxygen, not requiring ongoing medical care | 0 | 0 | 0 |
| 7. Not hospitalized, limitations on activity and/or requiring home oxygen | 0 | 0 | 0 |
| 8. Not hospitalized, no limitations on activities | 0 | 0 | 0 |
| Geographic Region |  |  |  |
| Argentina | 56 (29.9%) | 59 (31.9%) | 115 (30.9%) |
| Brazil | 54 (28.9%) | 54 (29.2%) | 108 (29.0%) |
| Mexico | 15 (8.0%) | 16 (8.6%) | 31 (8.3%) |
| Peru | 62 (33.2%) | 56 (30.3%) | 118 (31.7%) |
| Days from Symptom Onset* | -11.1 (3.52) | -11.2 (3.84) | -11.1 (3.68) |
| Body Mass Index (kg/m^2^)* | 29.4 (5.11) | 30.0 (5.29) | 29.7 (5.20) |
| **Comorbidities** |  |  |  |
| Hypertension | 50 (26.7%) | 50 (27.0%) | 100 (26.9%) |
| Obesity (BMI>=30 kg/m^2^) | 68 (36.4%) | 89 (48.1%) | 157 (42.2%) |
| Diabetes Mellitus | 31 (16.6%) | 33 (17.8%) | 64 (17.2%) |
| Coronary Artery Disease | 6 (3.2%) | 3 (1.6%) | 9 (2.4%) |
| History of Heart Failure | 2 (1.1%) | 2 (1.1%) | 4 (1.1%) |
| History of Cancer | 4 (2.1%) | 4 (2.2%) | 8 (2.2%) |
| Asthma | 6 (3.2%) | 10 (5.4%) | 16 (4.3%) |
| Chronic Obstructive Pulmonary Disease | 1 (0.5%) | 6 (3.2%) | 7 (1.9%) |
| Tuberculosis | 1 (0.5%) | 2 (1.1%) | 3 (0.8%) |
| HIV/AIDS | 1 (0.5%) | 0 (0.0%) | 1 (0.3%) |
| Severe Liver Disease | 1 (0.5%) | 0 (0.0%) | 1 (0.3%) |
| Severe Kidney Disease | 0 (0.0%) | 1 (0.5%) | 1 (0.3%) |

 * Displayed as Mean (Std. Dev.)

^†^Disease severity at baseline calculated as moderate disease = ordinal 5 + ordinal 4, severe disease = ordinal 3 + ordinal 2

### **Table S4. Additional secondary endpoints (modified intent-to-treat population)**

|  | **Infliximab** | **Shared Placebo** | | **Infliximab versus Shared Placebo** | |
| --- | --- | --- | --- | --- | --- |
|  | **Proportion of participants** | |  | **OR Estimate**  **(95% CI)** | ***p* value** |
| **Mortality^1^ at day 14** | 29/517 (5.6%) | 42/516 (8.1%) | | 0.627 (0.364, 1.080) | 0.0923 |
| **Mortality at Day 28** | 52/517 (10.1%) | 75/516 (14.5%) | | 0.593 (0.390, 0.904) | 0.0150 |
| **Clinical status^2^ at day 14 (N)** | 502 | 501 | | 1.318 (1.047, 1.659) | 0.0187 |
| **Clinical status at day 28 (N)** | 492 | 495 | | 1.448 (1.136, 1.846) | 0.0028 |

^1^ Mortality at 14 and 28 days calculated as odds of dying using logistic regression. A number less than 1 favors Infliximab.

^2^ Clinical status at 14 and 28 days calculated as proportional odds model by ordinal logistic regression. A number greater than 1 favors Infliximab.

### **Table S5. Demographics and baseline characteristics (intent-to-treat)**

| **Demographics and baseline characteristics** | **Infliximab**    **(N=531)** | **Shared Placebo**    **(N=530)** | **Total**    **(N=1061)** |
| --- | --- | --- | --- |
| Age in years* | 54.7 (14.87) | 54.9 (14.60) | 54.8 (14.73) |
| Male sex | 333 (62.7%) | 304 (57.4%) | 637 (60.0%) |
| Race |  |  |  |
| White | 328 (61.8%) | 339 (64.0%) | 667 (62.9%) |
| Black or African American | 80 (15.1%) | 73 (13.8%) | 153 (14.4%) |
| American Indian or Alaska Native | 5 (0.9%) | 5 (0.9%) | 10 (0.9%) |
| Asian | 12 (2.3%) | 17 (3.2%) | 29 (2.7%) |
| Native Hawaiian or Other Pacific Islander | 1 (0.2%) | 0 (0.0%) | 1 (0.1%) |
| Multiracial | 2 (0.4%) | 2 (0.4%) | 4 (0.4%) |
| Other | 76 (14.3%) | 71 (13.4%) | 147 (13.9%) |
| Unknown | 27 (5.1%) | 23 (4.3%) | 50 (4.7%) |
| Hispanic or Latino Ethnicity | 257 (48.4%) | 256 (48.3%) | 513 (48.4%) |
| Disease severity at baseline^†^ |  |  |  |
| Severe disease | 232 (43.7%) | 231 (43.6%) | 463 (43.7%) |
| Moderate disease | 299 (56.3%) | 299 (56.4%) | 598 (56.3%) |
| Clinical status (8-point ordinal scale) at baseline |  |  |  |
| 1. Death | 0 | 0 | 0 |
| 2. Hospitalized, on invasive ventilation or ECMO | 58 (10.9%) | 53 (10.0%) | 111 (10.5%) |
| 3. Hospitalized, on non-invasive ventilation or high flow oxygen devices | 174 (32.8%) | 178 (33.6%) | 352 (33.2%) |
| 4. Hospitalized, requiring supplemental oxygen | 275 (51.8%) | 279 (52.6%) | 554 (52.2%) |
| 5. Hospitalized, not requiring supplement oxygen, requiring ongoing medical care | 24 (4.5%) | 20 (3.8%) | 44 (4.1%) |
| 6. Hospitalized, not requiring supplement oxygen, not requiring ongoing medical care | 0 | 0 | 0 |
| 7. Not hospitalized, limitations on activity and/or requiring home oxygen | 0 | 0 | 0 |
| 8. Not hospitalized, no limitations on activities | 0 | 0 | 0 |
| Geographic Region |  |  |  |
| Argentina | 56 (10.5%) | 62 (11.7%) | 118 (11.1%) |
| Brazil | 54 (10.2%) | 55 (10.4%) | 109 (10.3%) |
| Mexico | 15 (2.8%) | 16 (3.0%) | 31 (2.9%) |
| Peru | 62 (11.7%) | 57 (10.8%) | 119 (11.2%) |
| USA - Northeast | 127 (23.9%) | 121 (22.8%) | 248 (23.4%) |
| USA - Midwest | 81 (15.3%) | 80 (15.1%) | 161 (15.2%) |
| USA - South | 98 (18.5%) | 97 (18.3%) | 195 (18.4%) |
| USA - West | 38 (7.2%) | 42 (7.9%) | 80 (7.5%) |
| Days from Symptom Onset* | -9.9 (4.38) | -9.9 (5.56) | -9.9 (5.00) |
| Body Mass Index (kg/m^2^)* | 32.1 (7.93) | 32.7 (8.11) | 32.4 (8.03) |
| **Comorbidities** |  |  |  |
| Hypertension | 211 (39.7%) | 215 (40.6%) | 426 (40.2%) |
| Obesity (BMI>=30 kg/m^2^) | 279 / 521 (53.6%) | 309 / 516 (59.9%) | 588 / 1037 (56.7%) |
| Diabetes Mellitus | 142 (26.7%) | 148 (27.9%) | 290 (27.3%) |
| Coronary Artery Disease | 38 (7.2%) | 27 (5.1%) | 65 (6.1%) |
| History of Heart Failure | 16 (3.0%) | 15 (2.8%) | 31 (2.9%) |
| History of Cancer | 35 (6.6%) | 34 (6.4%) | 69 (6.5%) |
| Asthma | 37 (7.0%) | 55 (10.4%) | 92 (8.7%) |
| Chronic Obstructive Pulmonary Disease | 24 (4.5%) | 26 (4.9%) | 50 (4.7%) |
| Tuberculosis | 3 (0.6%) | 4 (0.8%) | 7 (0.7%) |
| HIV/AIDS | 2 (0.4%) | 4 (0.8%) | 6 (0.6%) |
| Severe Liver Disease | 3 (0.6%) | 2 (0.4%) | 5 (0.5%) |
| Severe Kidney Disease | 3 (0.6%) | 7 (1.3%) | 10 (0.9%) |
| **Concomitant medication** |  |  |  |
| Remdesivir (Day 1 - Day 5) | 496 (93.4%) | 499 (94.2%) | 995 (93.8%) |
| Corticosteroids (Day 1 - Day 5) | 480 (90.4%) | 492 (92.8%) | 972 (91.6%) |
| Tocilizumab (any time post randomization) | 9 (1.7%) | 13 (2.5%) | 22 (2.1%) |
| Baricitinib (any time post randomization) | 6 (1.1%) | 12 (2.3%) | 18 (1.7%) |

 * Displayed as Mean (Std. Dev.)

^†^Disease severity at baseline calculated as moderate disease = ordinal 5 + ordinal 4, severe disease = ordinal 3 + ordinal 2

### **Table S6. Primary and secondary endpoints (intent-to-treat population)**

|  | **Infliximab** | **Shared Placebo** | **Infliximab versus Shared Placebo** | |
| --- | --- | --- | --- | --- |
|  | **Proportion of participants** | | **Estimate (95% CI)** | ***p* value** |
| **Recovery through day 28** | 421/531 (79.3%) | 405/530 (76.4%) | 1.122 (0.987, 1.275) | 0.0793 |
| **Mortality at day 14** | 29/531 (5.5%) | 42/530 (7.9%) | 0.632 (0.367, 1.090) | 0.0988 |
| **Mortality at day 28** | 53/531 (10.0%) | 75/530 (14.2%) | 0.614 (0.403, 0.935) | 0.0229 |
| **Clinical status at day 14 (N)** | 506 | 506 | 1.309 (1.041, 1.647) | 0.0213 |
| **Clinical status at day 28 (N)** | 496 | 500 | 1.435 (1.129, 1.825) | 0.0032 |

Estimates represent Recovery rate ratio, RRR (time to recovery) and Odds ratio, OR (mortality and clinical status)

### **Figure S1. Consort diagram**

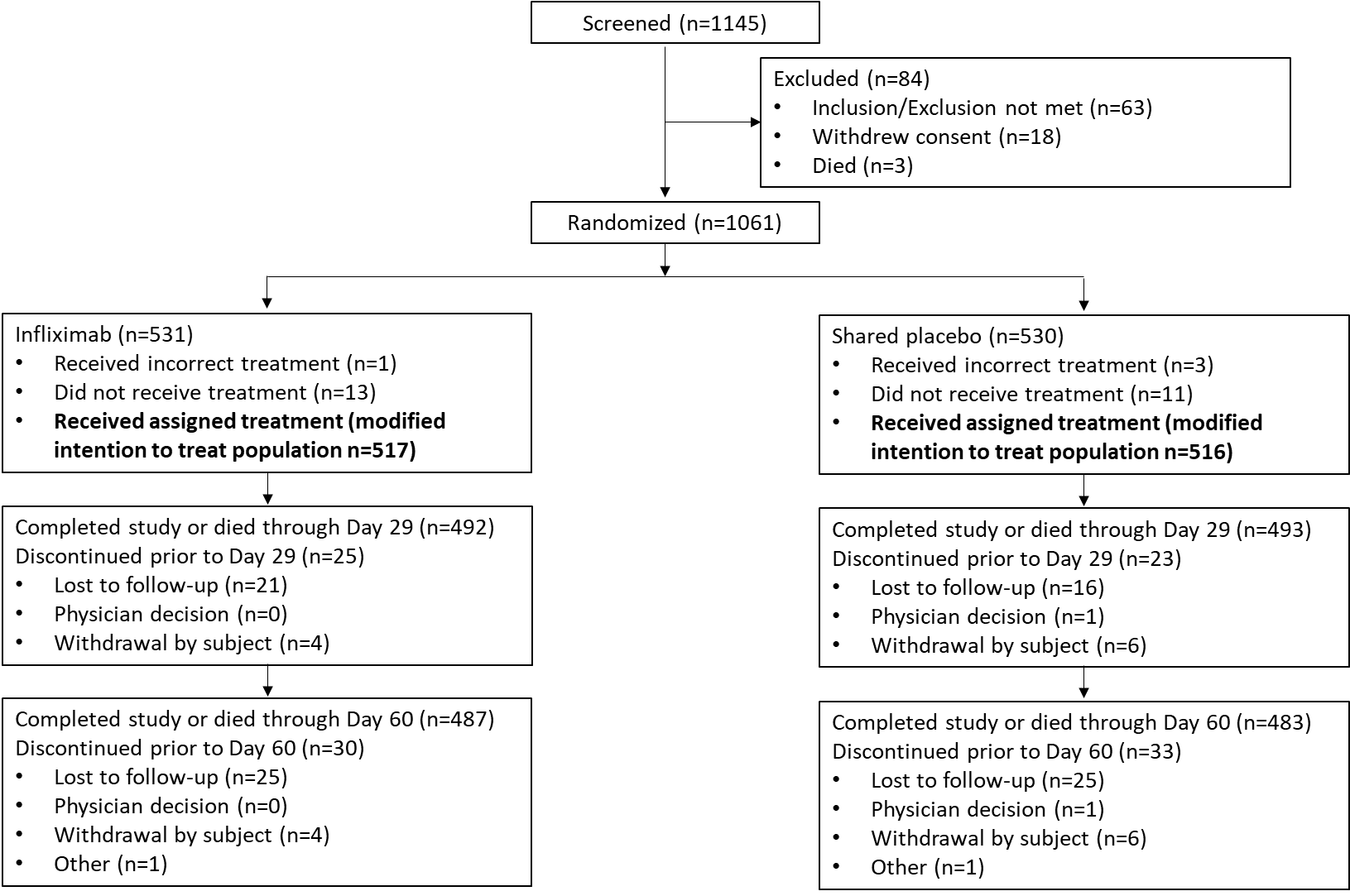

### **Figure S2. Forest plot of recovery rate ratios of time to recovery through day 28 by subgroup (modified intent-to-treat population)**

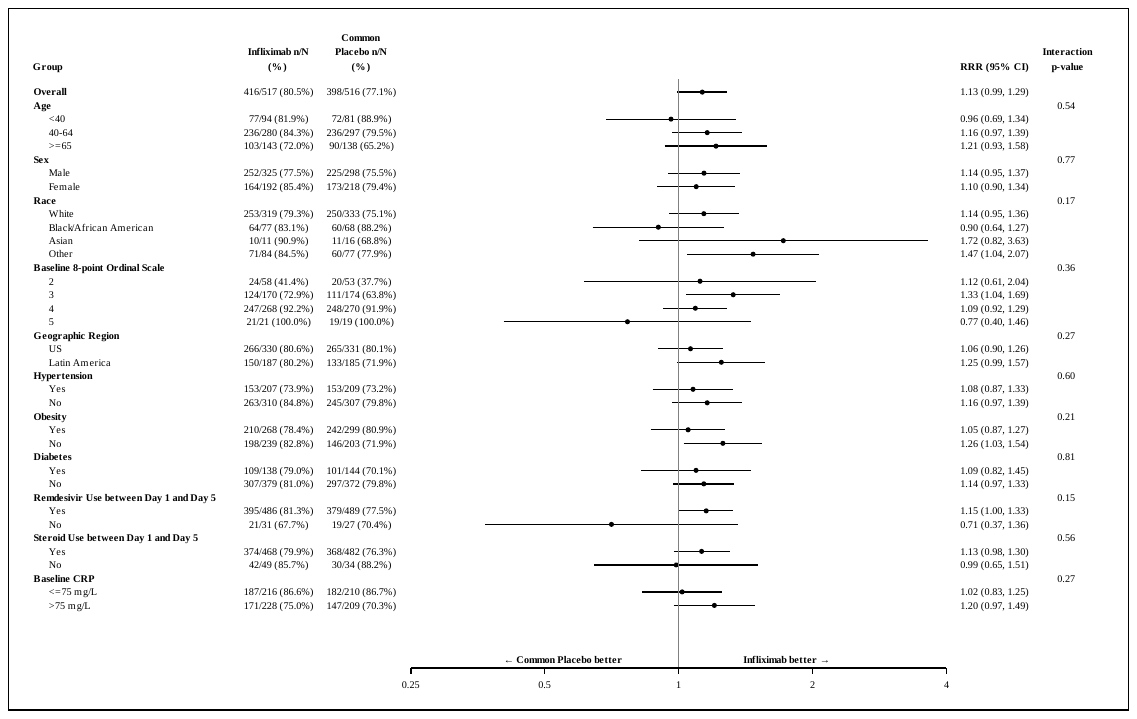

### **Figure S3. Forest plot of proportional odds model of clinical status score at day 14 by subgroup (modified intent-to-treat population)**

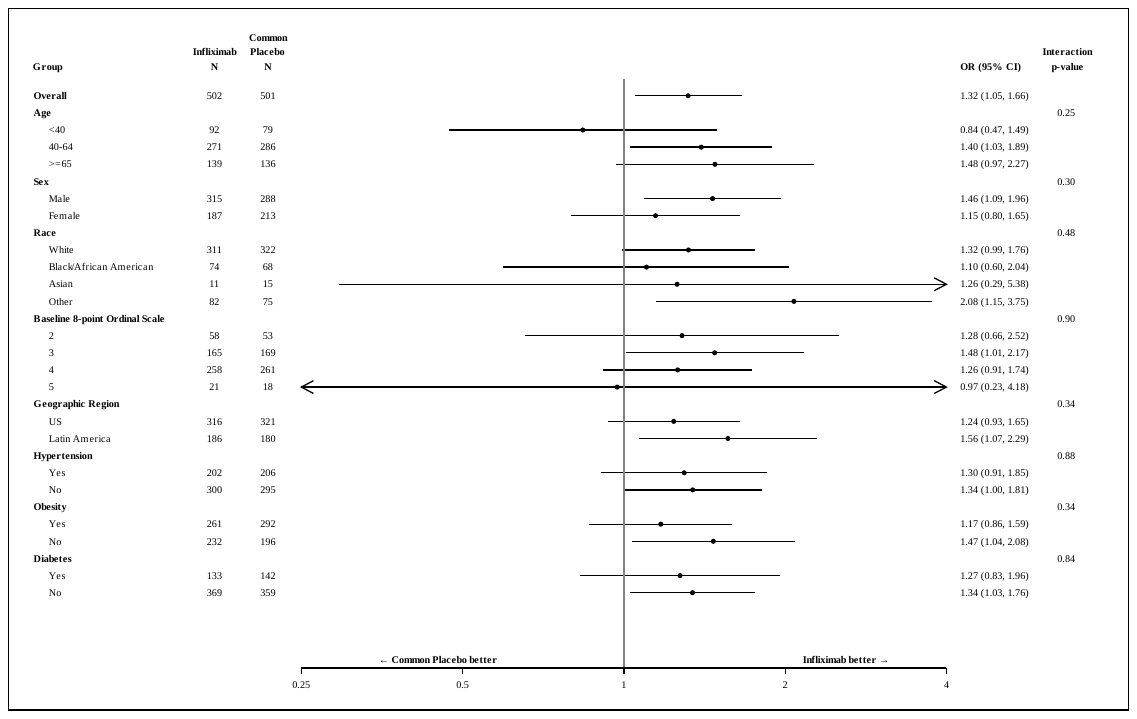

1.	Beigel JH, Tomashek KM, Dodd LE, et al. Remdesivir for the Treatment of Covid-19 - Final Report. N Engl J Med 2020;383:1813-26.
